# A family-aware All of Us benchmark for polygenic prediction and genomic machine learning

**DOI:** 10.64898/2026.10.22.26366229

**Authors:** Greycen Ren, Kaiden Wu, Christina Ren, Irura Nyiha, Kendall Wu, John Yang

## Abstract

Polygenic risk scores (PRSs) have improved substantially in predictive performance, yet results remain difficult to compare across studies that differ in cohorts, phenotype definitions, and evaluation procedures. Comparison to machine-learning methods is harder still, since their performance depends strongly on how data are partitioned and whether related individuals leak across splits. Here, we introduce a family-aware benchmark for polygenic prediction built on version 9 of the All of Us Research Program (AoU). The benchmark assigns entire family components to fixed discovery, calibration, and test partitions, reserves first-degree relatives for within-family evaluation, and separates models trained only on AoU discovery data from those using external GWAS summary statistics. This enables direct comparison between conventional PRS pipelines and machine-learning models trained under identical data budgets. We report results for 14 binary disease traits. Our strongest scores explain 5.7–24.9% of liability-scale variance, with particularly strong prediction for Alzheimer’s disease, type 2 diabetes, prostate cancer, glaucoma, and inflammatory bowel disease, and match or exceed published commercial and academic scores for most traits. Across 12 traits with interpretable within-family estimates of within-sibship-to-population effect ratio, 11 confidence intervals included a ratio of 1. Type 2 diabetes showed an attenuation ratio of 0.86 [0.76, 0.97].

## 1 Introduction

Polygenic risk scores (PRSs) have become increasingly predictive, driven by larger genome-wide association studies (GWASs) [Uffelmann et al., 2021] and improved statistical methods [Prive et al., 2020, Zheng et al., 2024, Park and Chung, 2026, Williams et al., 2026]. However, reported performance is difficult to compare across studies. Most PRS methods are trained on one or more large external GWASs and evaluated in an independent cohort [Ge et al., 2019, Lloyd-Jones et al., 2019, Zheng et al., 2024], so differences in methodology are confounded with differences in discovery power, phenotype definitions, and evaluation procedures. This paradigm is appropriate for validating a fixed clinical score, but less suited for developing and comparing new methods.

Comparison to machine-learning methods is harder still. Their performance depends strongly on how data are partitioned, and can be inflated when related individuals are shared across training and evaluation sets [Jeong et al., 2025, Ellis et al., 2024]. Efforts to standardize PRS reporting [Wand et al., 2021, Lambert et al., 2021] and benchmarks of statistical methods in the UK Biobank [Sudlow et al., 2015, Park and Chung, 2026, Pain et al., 2021, Chen et al., 2026] have begun to address these issues, and recent work has proposed the All of Us Research Program (AoU) [The All of Us Research Program Genomics Investigators, 2024] as a resource for PRS discovery and validation [Tsuo et al., 2026, Williams et al., 2026, Das and Cui, 2026, Wu et al., 2026]. However, these works have yet to provide a reusable framework with fixed, leakage-safe data partitions that supports both internal-discovery-only and externally powered discovery, and that evaluates prediction among unrelated individuals across ancestries [Kachuri et al., 2024], and within families, where shared ancestry and family-level confounding are substantially reduced [Selzam et al., 2019, Howe et al., 2022, Young et al., 2022]. Within-family prediction is also the most relevant evaluation for reproductive applications such as embryo selection, where scores are used to compare siblings. In this work, we make two key contributions: (1) we construct a family-aware benchmark in AoU v9 that assigns entire family components to fixed discovery, calibration, and test partitions and reserves first-degree relatives for within-family evaluation, and (2) we define two modeling tracks, AoU internal-discovery-only and external-permitted, so that conventional PRS pipelines and machine-learning models can be compared under identical data budgets. We then populate the benchmark with baselines for 14 binary disease traits and report population, within-family, and cross-ancestry performance. Published scores from Herasight [Moore et al., 2025], Nucleus [Cordogan et al., 2025], and academic groups [Thompson et al., 2024, Mars et al., 2022] are included as context rather than as formal comparisons.

We structure the paper as follows. Section 2 describes the benchmark design and modeling tracks. Section 3 details data processing, partitioning, phenotype definitions, and evaluation metrics. Section 4 presents results for population prediction, within-family attenuation, and cross-ancestry transport, and Section 5 discusses implications and limitations. We summarize our main findings below.

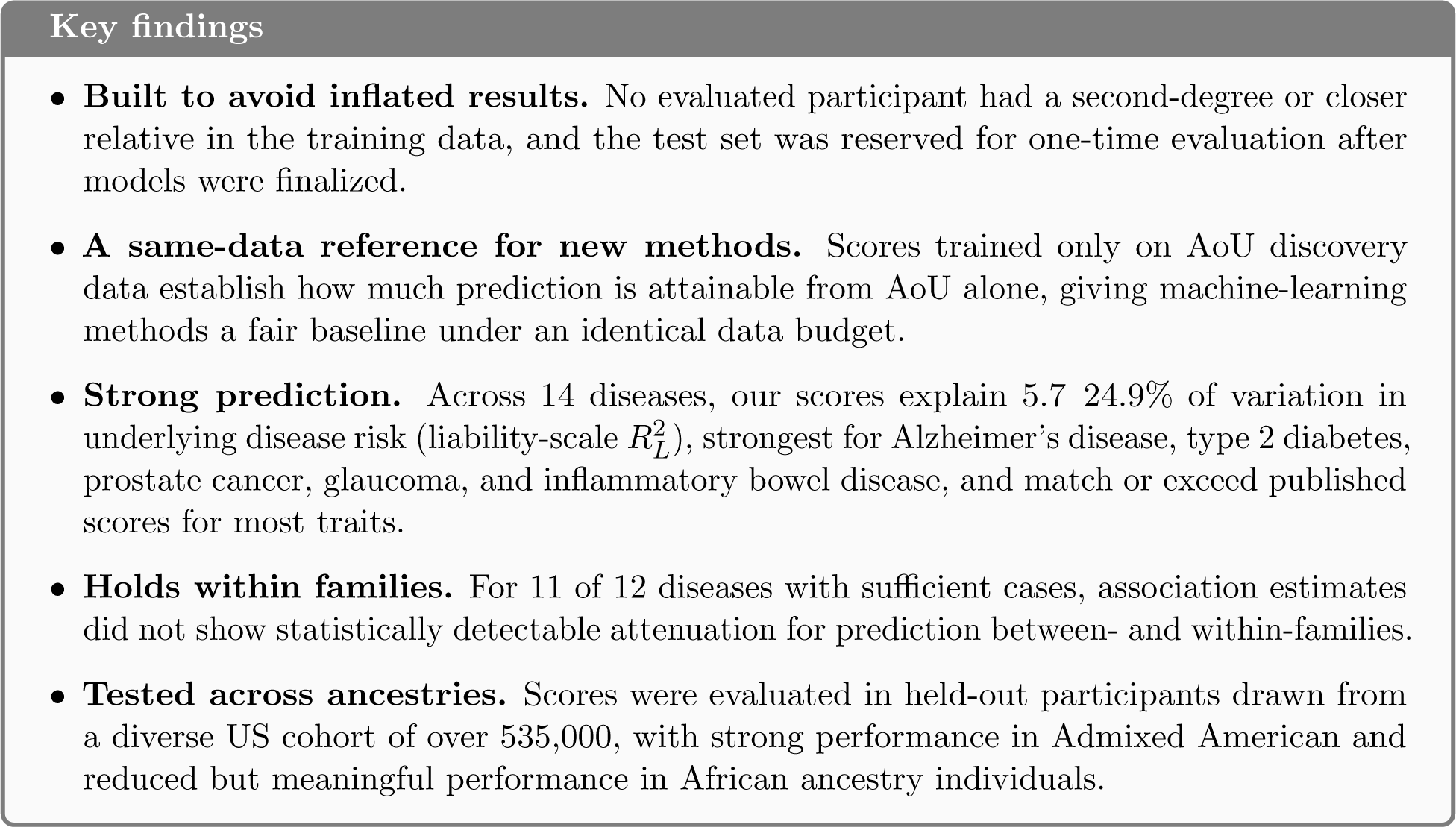

## 2 Benchmark design

### 2.1 Design goals

The benchmark was designed around four goals. First, it should allow iterative discovery using individual-level AoU data without using the final evaluation sample for model selection. Second, it should prevent obvious relatedness leakage by assigning connected family components, rather than nominally unrelated individuals, to data partitions. Third, it should support models trained from different information sources under explicitly labeled tracks. Fourth, it should preserve evaluation layers that answer different scientific questions rather than reducing all model performance to a single headline metric.

This led to a benchmark architecture with distinct roles for discovery, calibration, evaluation, and family-based confirmation. The discovery partition is the only AoU sample from which phenotype-associated training signal may be learned. The calibration partition is used for operations such as hyperparameter selection, component-score weighting, or post-hoc scaling that are explicitly permitted by the model specification. A separate evaluation sample is preserved for one-time validation after a model and analysis plan are frozen. Family-based confirmation is treated as an additional validation axis rather than folded into the ordinary population test set.

This separation was intentionally conducive to *internal-discovery-only training*: new methods can use the AoU discovery genomes and phenotypes aggressively, for GWAS, feature selection, representation learning, interaction discovery, rare-variant aggregation, or direct prediction, while the provenance of every reported result remains clear. Repeated method development can therefore occur without silently converting the full biobank into training data.

### 2.2 Family-aware partitioning

AoU participants were organized using a genome-wide relatedness graph before the primary modeling partitions were created. Family components were constructed across ancestries to enable cross-ancestry familial structures for admixed populations and minimize cross-ancestry genetic leakage. First-degree relatives and their entire family components were first isolated as the downstream within-family holdout. Remaining individuals were allocated component-wise across the remaining three roles. We elaborate more on this in Section 3.2

### 2.3 Separate model-development tracks

We distinguish separate benchmark tracks according to where phenotype-associated training information originates in Table 1. This distinction is important because an externally trained PRS and a model learned from AoU individual-level data solve different statistical problems even when validated in the same evaluation sample.

**Table 1:** Primary model-development tracks in the Reticular AoU benchmark. The two model-development tracks emphasize different scales of discovery information for phenotype-associated signal. Reference resources that do not contain target-phenotype labels, such as annotation databases or LD resources, are governed separately by each model specification.

| Track | Phenotype-associated training signal | Primary purpose |
| --- | --- | --- |
| External-permitted | Any clean GWAS or externally-trained model weights | Assess validation of strongest PRSs and scaling of summary statistic-powered methods |
| AoU internal-discovery-only | Discovery for learning; calibration for pre-specified tuning | Benchmark AoU-only training signal for method development |

The external-permitted track anchors the benchmark to the strongest summary statistic-based predictors currently available, eligible under an institutional-level exclusion rule. We consider any GWAS that does not directly incorporate data from explicit contribution partners to AoU at an institutional-or biobank-level as eligible. Hence, large-scale American biobanks including the Million Veterans Program (MVP) [Verma et al., 2024] are considered eligible, even though we cannot extensively audit individual-level overlap. This characterization is generally accepted by previous studies that have developed PRS methods in MVP and validated in AoU [Haydarlou et al., 2026, Smith et al., 2026]. The AoU internal-discovery-only track was designed as a fair comparison for downstream machine learning and method development: it defines a separate task in which the target phenotype is learned only from the discovery partition, making it possible to compare conventional GWAS-plus-PRS pipelines with methods that operate directly on individual-level genomes.

### 2.4 Evaluation layers

The benchmark reports several evaluation layers rather than a single leaderboard. Population prediction evaluation measures out-of-sample disease association and the variance explained on a liability-scale. Cross-ancestry evaluation measures determine whether a model trained in one ancestry or jointly across ancestries transports well to other ancestries. Within-family evaluation measures whether the same score remains predictive even when the contrast is driven by genetic segregation within a family. These layers are deliberately separated, as a model may improve populution prediction metrics while worsening cross-ancestry transfer or maintain ancestry performance while losing signal within families. Reporting outcomes separately creates a more informative target for future method development and validation than maximizing a single aggregated metric. For this reason, the benchmark should be viewed as an experimental infrastructure rather than a static leaderboard. New models can be added over time while the phenotype definitions, split assignments, evaluation code, and baseline results remain versioned. For reproducibility of our existing results, we provide phenotype definitions in Table B1 and detail our evaluation settings in Section 3.7. We cannot export exact split assignments with participant IDs from AoU, but similar split generation protocols should yield comparable results.

## 3 Methods

### 3.1 All of Us Data

All of our analyses used the AoU Research Program Controlled Tier dataset version 9 [The All of Us Research Program Genomics Investigators, 2024] for whole-genome sequencing and longitudinal EHR-derived phenotypes for over 535,000 participants in a diverse American cohort, including large contributions from European (EUR), African (AFR), and Admixed American (AMR) ancestry cohorts. Existing data also include individuals of East Asian (EAS), South Asian (SAS), and Middle Eastern (MID) descent. Data were accessed through the Researcher Workbench under Controlled Tier (C2025Q4R6). Ancestry was determined from the AoU-provided Echo v4 r2 ancestry-prediction table. We took the Baseline-LD annotation set [Gazal et al., 2017, Márquez-Luna et al., 2021] and ran variant-level quality control, keeping only variants with non-zero variance and call rate ≥ 0.98 with an adaptive minor allele count (MAC) per ancestry, with a floor of 20 and a ceiling of 100, otherwise using a cutoff at 1% of the called cohort. This maintained a MAC 100 threshold for EUR, while for smaller ancestries, the cutoff was adaptive to include more variants, down to a floor of 20. No Hardy-Weinberg filter was applied, leaving us with a total of 7,724,164 biallelic variants. Principal components were fit exclusively inside the discovery train set, sampling up to 10,000 individuals per ancestry for diversity and balance, before projecting onto remaining individuals. Linkage disequilibrium (LD) was estimated within the EUR ancestry discovery cohort on a set of unrelated individuals.

### 3.2 Relatedness graph and partition algorithm

We constructed a genome-wide relatedness graph in which participants are vertices and edges connect individuals whose estimated kinship exceeds the benchmark threshold. We utilize the KING robust estimator [Manichaikul et al., 2010] to compute kinship coefficients, building kinship edges at the second-degree relative threshold with kinship coefficient ≥ 0.0884. Connected components, rather than individual participants, formed the atomic unit for primary split assignment. The first-degree within-family holdout set was constructed first, with entire components held out before the remaining graph was partitioned. Individuals were then assigned component-wise, allocating to the discovery, calibration, and test partitions at a 70/5/25 ratio. The number of individuals by split and ancestry is shown below in Table 2.

**Table 2:** Sample allocation by ancestry across discovery, calibration, locked test, and within-family holdout sets. The 70/5/25 split was applied only after the 58,726-member family holdout was removed.

| Ancestry | Discovery | Calibration | Locked test | Within-family holdout | Total |
| --- | --- | --- | --- | --- | --- |
| AFR | 60,717 | 4,292 | 21,516 | 14,275 | 100,800 |
| AMR | 62,436 | 4,484 | 22,416 | 18,592 | 107,928 |
| EAS | 10,259 | 742 | 3,755 | 1,137 | 15,893 |
| EUR | 195,034 | 13,931 | 69,655 | 24,094 | 302,714 |
| MID | 1,370 | 102 | 484 | 195 | 2,151 |
| SAS | 4,039 | 296 | 1,408 | 433 | 6,176 |
| **Total** | **333,855** | **23,847** | **119,234** | **58,726** | **535,662** |

The resulting benchmark contains four logical roles: discovery, calibration, test, and family confirmation. The discovery partition may be used repeatedly for phenotype-associated feature learning. Calibration may be used only for pre-specified tuning operations or model selection, before evaluation is run on the test set. Family confirmation is used for within-family estimands and is not treated as an ordinary unrelated test sample.

Where ancestry-specific analyses were performed, the split assignment was inherited directly from the same global family-aware partition rather than reconstructed independently within each ancestry. As family components were constructed across ancestry, each split for each ancestry was clean with respect to the discovery-train of any other ancestry, and ancestry-specific splits were strict subsets of global splits.

### 3.3 Phenotype definitions

Binary phenotypes were defined from AoU EHR data using trait-specific OMOP concept sets and eligibility rules. Where clinically appropriate, case definitions required repeated diagnosis dates separated in time to reduce single-code misclassification. Control definitions incorporated trait-specific age and observation requirements and excluded related or precursor diagnoses when necessary. Sex-specific cancers are restricted to the relevant eligible population. Specific phenotype definitions for the 14 included binary traits are detailed in Table B1, with additional specific concept IDs further detailed in Appendix D.

### 3.4 Benchmark track definitions

A model is labeled *external-permitted* when any phenotype-associated training signal is obtained from sources outside AoU. AoU internal discovery and phenotype-associated training signal and AoU calibration are permitted under this framework as well. A model is labeled *AoU internal-discovery-only* when all target-phenotype-associated training signal is estimated from individuals in the discovery partition. The use of external LD references, functional annotations, genome annotations, or unsupervised reference data does not by itself change the training-track label, provided these resources do not contain target-phenotype labels from AoU evaluation participants. Internally built LD resources and PCA projections should be fit only on the discovery set and should not include any individuals in the calibration, test, or within-family holdout, regardless of benchmark track.

### 3.5 External-permitted PRS benchmarks

For each trait, we identified clean, large external GWAS summary-statistic datasets with phenotype definitions sufficiently close to the AoU target phenotype. To account for misspecification in external GWAS sources [Prive et al., 2022], external summary statistics were harmonized to the benchmark variant panel by rsID with strand and allele-frequency–assisted palindrome resolution, retaining only biallelic SNPs with valid effect estimates. Palindromic variants were resolved by frequency when both source and panel MAF ≤ 0.4 and otherwise excluded. When several independent sources were used, they were aggregated via meta-analysis. Specific provenance for external consortia and meta-analyses are documented in Table B5. Where available, publicly available GWAS from UK Biobank [Sudlow et al., 2015], FinnGen[Kurki et al., 2023], and MVP[Verma et al., 2024] contributed to final meta-analysis. Internal AoU GWAS was run on the discovery cohort and contributed to the meta-analysis. Case and control counts for the final meta-analyses are provided in Table B3. We utilized SBayesRC, an LD-aware Bayesian shrinkage methodology that leverages functional annotations to convert GWAS summary statistics into a posterior effect estimator [Zheng et al., 2024]. This was fitted across the ∼ 7.7M SNP panel. We followed the settings in the original paper, matching the 591-block LD eigen-decomposition (which we refit on our discovery cohort), SNP-level QC, annotations, and MCMC settings.

### 3.6 AoU internal-discovery-only baseline construction

Internal-discovery-only baselines were trained using target phenotypes from the discovery partition. In this initial benchmark, we utilized SAIGE [Zhou et al., 2018] to fit discovery GWAS for each trait. SAIGE was selected due to strong binary-trait calibration for imbalanced phenotypes. Discovery case and control counts are provided in Table B2. Discovery GWAS marginal effect estimates were then harmonized to the variant panel and fed into SBayesRC to yield a posterior effect estimator. These internal-discovery-only benchmarks are such that downstream methods are compared with a strong statistical baseline under the same data budget. Due to discovery cohort size, we focused primarily on EUR-ancestry GWAS and provide evaluation across ancestry cohorts. From cross-ancestry comparisons, we used score-only AUC as the primary metric to avoid reliance on uncertain ancestry-specific disease prevalence assumptions. When a model required hyperparameter selection, these were selected in calibration or family-aware cross-validation confined to discovery. The final fitted models are then evaluated without refitting on the evaluation sample.

### 3.7 Evaluation Metrics

We evaluated PRSs in two tracks for both predictive and explanatory power. In the predictive setting, we computed predictive metrics on the test set and report score-only AUC. Our calibration set design permits a separate covariate-adjusted predictive performance, where the covariates and PRS are fit in calibration, frozen, and then evaluated on the test set. In the explanatory setting, we followed the methodology in Moore et al. [2025], isolating the incremental contribution of a score by residualizing the PRS against nuisance covariates

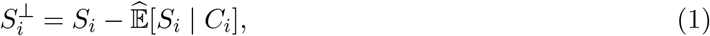

and estimated the observed-scale variance explained from the pre-specified outcome model. We residualized against age and the first 10 genetic principal components as covariates. The primary binary-trait metrics are liability-scale *R*^2^ and AUC. The conversion to liability-scale variance explained is detailed in Appendix A.1. Uncertainty was estimated through 2000 individual bootstrap replicates.

### 3.8 Within-family evaluation

We evaluated whether the PRS association attenuates within families on the first-degree within-family holdout. The within-sibship genetic effect was estimated with a within/between sibship decomposition that requires no parental genotype information. For each proband *i* in family *f* (*i*) the score was decomposed into

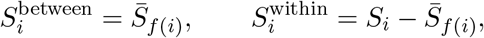

and, per trait, we fit

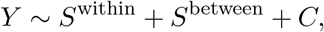

with covariates *C* = age and sex. The coefficient on *S*^within^ is identified solely by within-sibship score differences arising from Mendelian segregation; shared ancestry, shared environment, and all parental-score components are absorbed by *S*^between^. The population effect *β*_pop_ was estimated from *Y ∼ S*_proband_ + *C* on the same modeled rows, and within-family retention was summarized as

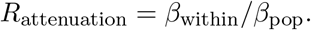

This design was chosen because parental genotypes are available for only a minority of holdout families. For more details on evaluation settings, refer to Appendix A.2. Parental-adjustment estimates restricted to families with at least one genotyped parent, together with per-stratum composition and estimates, are reported in Table C4.

Uncertainty was estimated by nonparametric bootstrap resampling of whole families (2,000 replicates): in each replicate both models were refit on the same resampled families and the ratio recomputed, preserving within-family dependence and the covariance between the two coefficients. Confidence intervals are empirical percentile intervals from the bootstrap distribution. Estimates were flagged non-interpretable when the *β*_pop_ interval included zero. Estimates made from 20 or fewer cases are suppressed.

### 3.9 Cross-ancestry evaluation

Ancestry-stratified analyses inherited the global family-aware split assignment. Scores were evaluated separately within ancestry groups using locally estimated nuisance coefficients under the frozen PC basis and evaluation specification. While not provided in this report, the benchmark formulation is easily compatible with multi-ancestry methods. For results across ancestries, metrics should report which benchmark track is followed, which ancestry groups contribute training signal, and distinguish cross-ancestry portability from joint multi-ancestry training.

### 3.10 Published comparator context

Published PRS results from Thompson et al. [Thompson et al., 2024], Mars et al. [Mars et al., 2022], Herasight [Moore et al., 2025], and Nucleus [Cordogan et al., 2025] are used as contextual references. When necessary, reported estimates are transformed into the benchmark metric under frozen trait-specific prevalence assumptions. These cross-study values are not used to select benchmark models and are not interpreted as formal pairwise tests because the underlying cohorts, phenotypes, training sources, and analysis procedures differ.

## 4 Results

### 4.1 Liability-scale variance explained across sources

We first characterized our meta-analyses for the 14 binary traits in terms of liability-scale variance explained relative to two other commercial groups in Herasight [Moore et al., 2025] and Nucleus [Cordogan et al., 2025], as well as two academic works [Thompson et al., 2024, Mars et al., 2022]. Note that due to different discovery cohorts, evaluation settings, data split methodologies, and phenotype definitions, these comparisons are not strictly head-to-head, but rather demonstrate that meta-analysis of publicly available GWAS yields strong polygenic risk prediction within AoU v9. These results are plotted in Figure 1.

**Figure 1:**
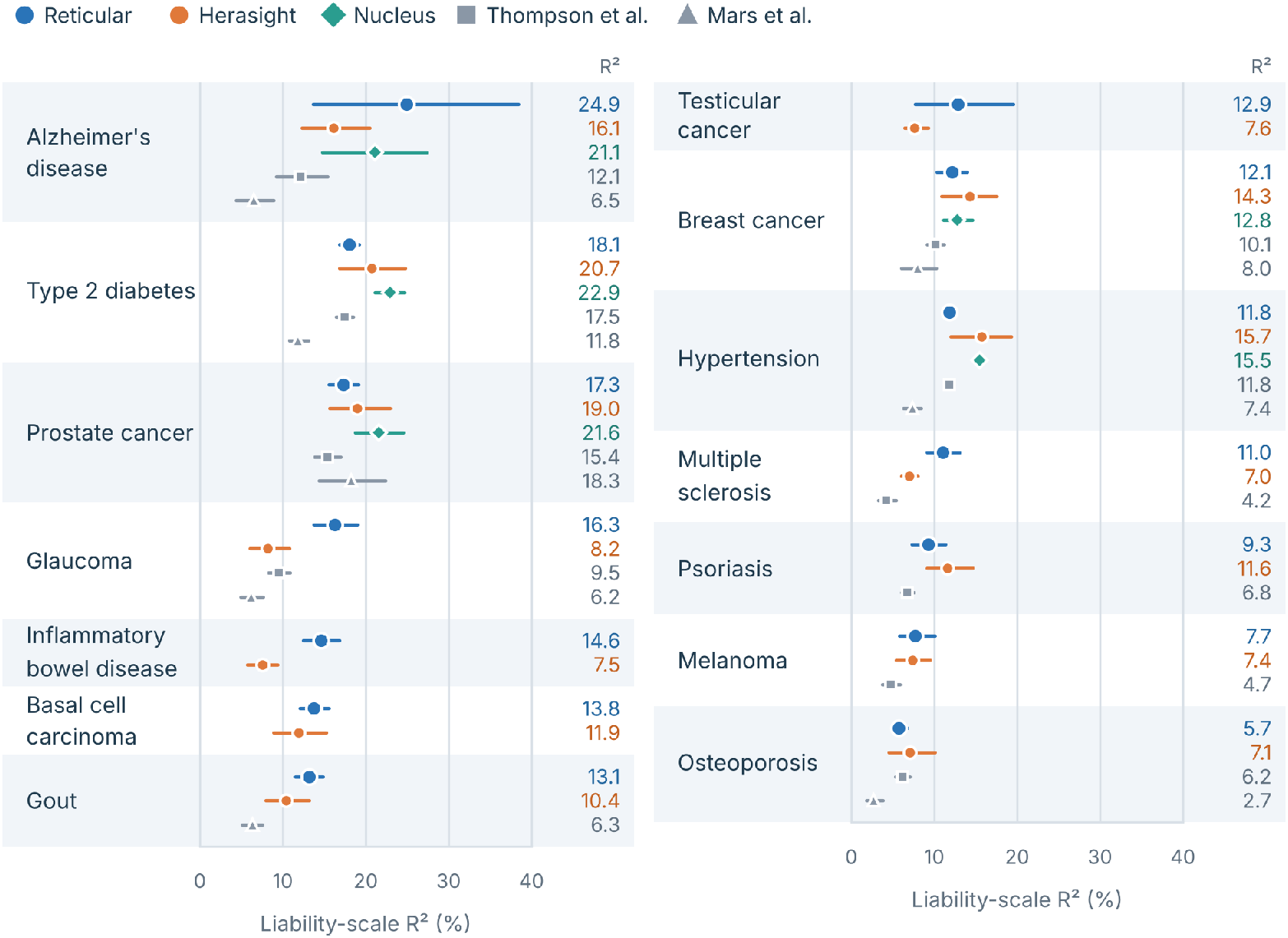
Liability-scale variance explained across 14 binary traits for external-permitted track on EUR ancestry individuals. Reticular refers to our meta-analysis of external and AoU internal GWAS. We exceed or match the liability variance explained for most traits, though exact phenotype definitions, evaluation cohorts, and populations differ by source. Numerical values provided in Table C1.

These meta-analysis results include external GWAS and internal GWAS sources. We also wanted to characterize the discovery power inherent within AoU through our family-aware data splitting mechanism. We were unable to run a meaningful internal-discovery-only GWAS on testicular cancer due to the low case count, so we demonstrate this across the remaining 13 binary traits. These results are displayed in Figure 2. Numerical results are provided in Table C1 and Table C2. External-permitted PRSs explain more variance in liability-scale than internal-discovery-only, which is unsurprising given the difference in discovery power. These results demonstrate the importance of separating raw metric strength from discovery power for fair comparisons in method development.

**Figure 2:**
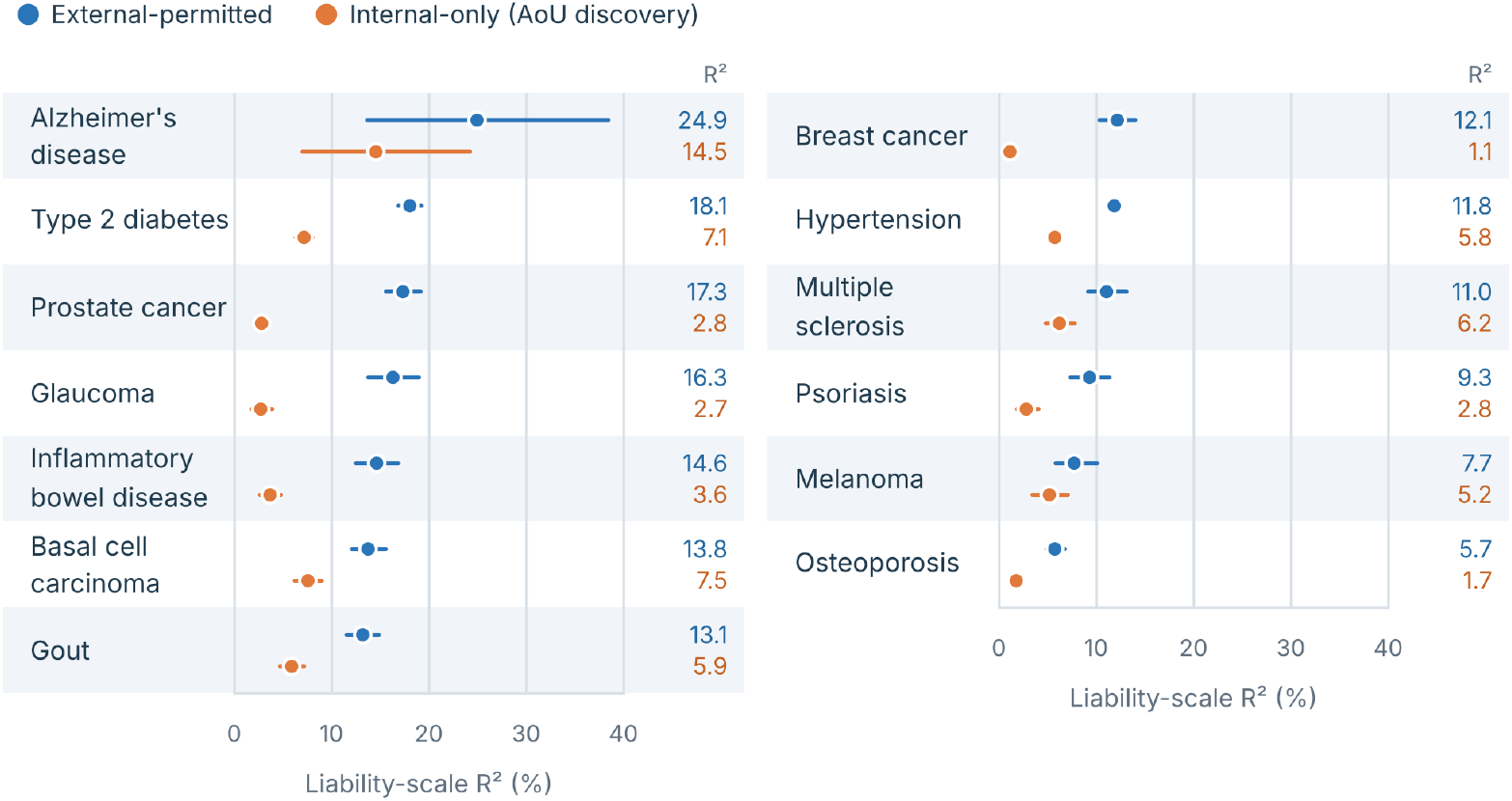
Comparison of liability-scale variance explained across 13 binary traits (testicular cancer dropped) between external-permitted and internal-discovery-only benchmark tracks on EUR ancestry individuals. Use of high-power external GWAS sources strongly strengthens performance of polygenic risk predictors. confidence intervals including 1. For type 2 diabetes, we observe slightly worse within-family than between-family performance (attenuation ratio 0.86 [0.76, 0.97]). Estimates for Alzheimer’s disease are unstable due to a limited case count in the within-family holdout. Estimates for testicular cancer are suppressed due to a limited case count.

### 4.2 Within-family analysis for external-permitted scores

We then hoped to characterize the attenuation of polygenic risk prediction within-family. Plots of the attenuation ratio, the ratio between the within-sibship and population effect, are shown in Figure 3. For 12 interpretable traits, we found that the predictive power of PRSs was not significantly different within-sibship from the population at large for 11 PRSs, with attenuation ratio

**Figure 3:**
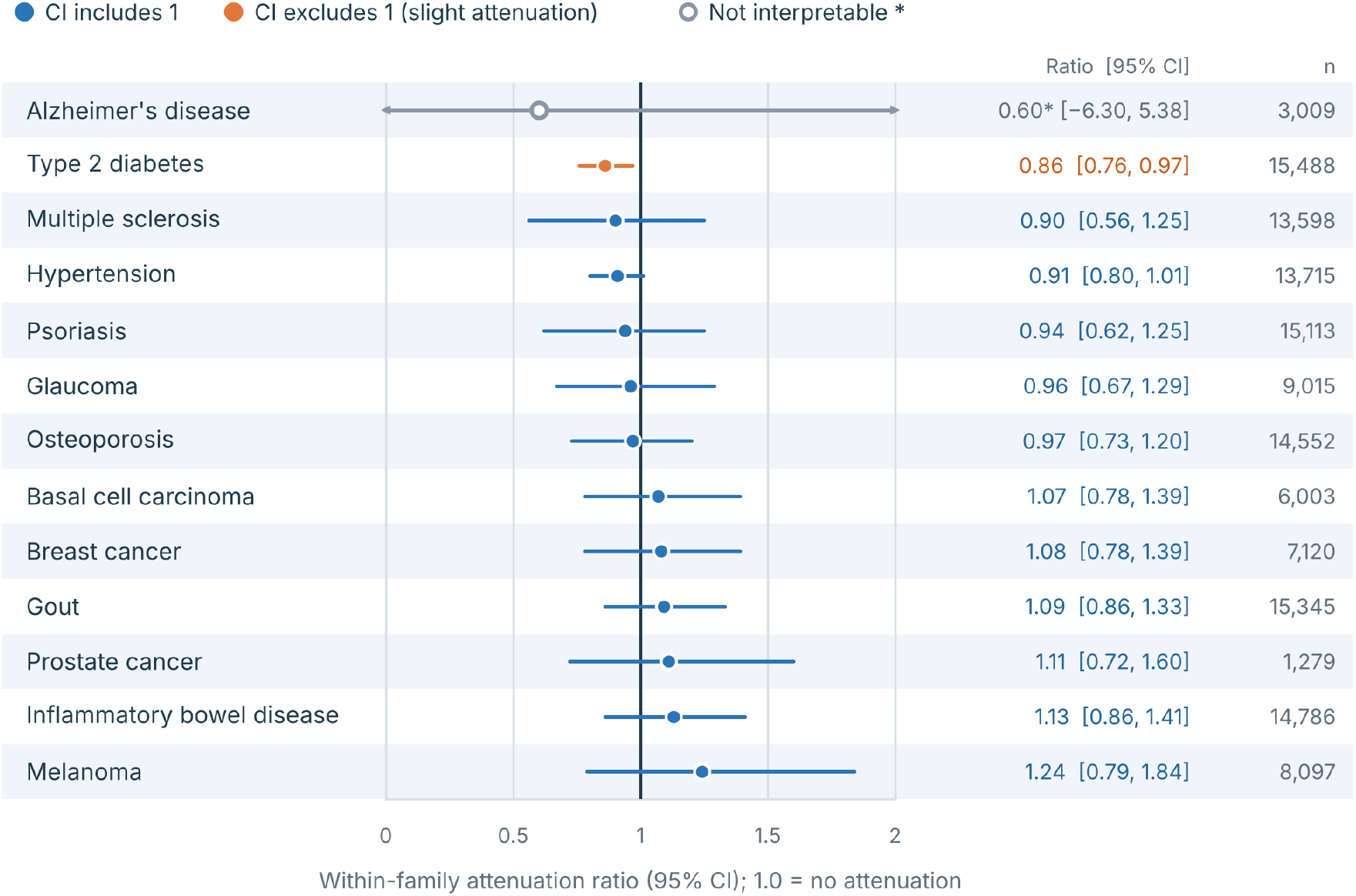
Within-family attenuation ratio across 13 binary traits for external-permitted PRSs. *n* refers to the number of probands modeled. Alzheimer’s disease estimates are unstable due to low case counts. Testicular cancer estimates are suppressed as per AoU policy. 11 binary traits do not meaningfully attenuate within family, with confidence intervals including 1, while type 2 diabetes shows slight attenuation in within-family evaluation. Numerical values are provided in Table C4.

### 4.3 Cross-ancestry transport for external and internal methods

We characterized cross-ancestry transport using score-only PRS AUC as our primary predictive metric. Unlike liability-scale variance explained, AUC does not require assumptions about ancestry-specific population disease prevalence, which are unavailable or uncertain for several trait–ancestry pairs. External-permitted PRS results are reported in Table 3, with internal-discovery-only results provided in Table C3. Following AoU data agreements, we suppressed estimates made from 20 or fewer cases. Among combinations with evaluable estimates in both groups, point-estimate performance was generally higher for AMR than AFR ancestry, except for breast cancer. Full case and population sizes for traits across ancestries are provided in Table B4.

**Table 3:** Cross-ancestry score-only AUC for external-permitted PRSs. 95% confidence intervals are computed on 2000-replicate stratified bootstraps. Score-only PRS AUC was computed on the entire test set. As per AoU data policy, estimates made from 20 or fewer cases are suppressed.

| Trait | EUR | AFR | AMR | EAS | MID | SAS |
| --- | --- | --- | --- | --- | --- | --- |
| Alzheimer’s disease | 0.690 [0.642, 0.734] | — | 0.754 [0.674, 0.823] | — | — | — |
| Basal cell carcinoma | 0.663 [0.652, 0.675] | — | 0.717 [0.644, 0.783] | — | — | — |
| Breast cancer | 0.662 [0.650, 0.675] | 0.623 [0.593, 0.653] | 0.617 [0.586, 0.649] | 0.641 [0.564, 0.711] | — | — |
| Glaucoma | 0.733 [0.714, 0.752] | 0.669 [0.635, 0.704] | 0.705 [0.657, 0.749] | 0.622 [0.519, 0.725] | — | — |
| Gout | 0.703 [0.690, 0.715] | 0.609 [0.583, 0.634] | 0.669 [0.630, 0.708] | 0.726 [0.675, 0.773] | — | — |
| Hypertension | 0.647 [0.642, 0.653] | 0.570 [0.559, 0.581] | 0.632 [0.619, 0.645] | 0.660 [0.629, 0.690] | 0.653 [0.568, 0.730] | 0.645 [0.582, 0.702] |
| Inflammatory bowel disease | 0.761 [0.744, 0.778] | 0.649 [0.589, 0.706] | 0.704 [0.650, 0.755] | — | — | — |
| Melanoma | 0.670 [0.648, 0.695] | — | — | — | — | — |
| Multiple sclerosis | 0.748 [0.729, 0.768] | 0.586 [0.533, 0.643] | 0.692 [0.628, 0.755] | — | — | — |
| Osteoporosis | 0.613 [0.603, 0.622] | 0.580 [0.548, 0.615] | 0.593 [0.563, 0.625] | 0.557 [0.495, 0.620] | — | — |
| Prostate cancer | 0.653 [0.643, 0.661] | 0.579 [0.556, 0.603] | 0.616 [0.588, 0.641] | 0.609 [0.548, 0.670] | 0.603 [0.461, 0.729] | 0.512 [0.396, 0.635] |
| Psoriasis | 0.694 [0.674, 0.711] | 0.540 [0.459, 0.624] | 0.647 [0.592, 0.699] | 0.653 [0.514, 0.791] | — | — |
| Testicular cancer | 0.775 [0.714, 0.833] | — | — | — | — | — |
| Type 2 diabetes | 0.699 [0.692, 0.707] | 0.582 [0.568, 0.594] | 0.649 [0.635, 0.662] | 0.638 [0.597, 0.678] | 0.611 [0.513, 0.700] | 0.613 [0.557, 0.671] |

For the subset of trait–ancestry combinations with suitable literature-derived lifetime-risk estimates and sufficient evaluation cases, we additionally computed liability-scale variance explained, shown in Figure 4, using lifetime risk estimates provided in Table B6. These estimates provide complementary information about predictive performance but are restricted to combinations with both suitable prevalence estimates and sufficient case counts; hence, they should not be interpreted as a comprehensive cross-ancestry comparison of liability-scale variance explanation.

## 5 Discussion

This work reframes PRS validation in AoU as a benchmark-design problem. Existing company whitepapers have importantly demonstrated that modern PRSs can reach substantial disease-prediction performance and can retain signal within families [Moore et al., 2025, Cordogan et al., 2025]. Our objective is somewhat complementary. While we are interested in disease-predictive performance of modern PRSs, we also recognize the need for a standardized evaluation setting in which existing PRSs can iteratively be seen as baselines for the next generation of genomic methods.

Our central methodological choice is a family-aware data partition. A random holdout is sufficient only when the final model is trained once and close relatives are absent. It is less satisfactory for an iterative research program in which new models are repeatedly proposed, individual-level AoU data are used for feature discovery, and family structure may create hidden dependence between samples. By freezing family-aware discovery, calibration, evaluation, and within-family partitions, we make the information boundary explicit. The same split can support conventional GWAS-plus-PRS pipelines and more flexible machine-learning models without redefining what constitutes an independent test.

The distinction between external-permitted and internal-discovery-only performance is also important. Large external GWASs provide far more association power than a single internal discovery split for many diseases, so an external PRS is an intentionally difficult baseline. Internal-discovery-only results provide an important measurement for the efficiency with which a model converts a fixed set of individual-level genomes and phenotypes into held-out prediction. This is the relevant quantity for assessing new methods that claim to learn more from the same biobank data. The most powerful methods can then be scaled to larger sizes and provide more powerful PRS prediction.

The nested nature of our data partitioning methodology across ancestries is also important for evaluation and development of multiancestry methods. Prior work has demonstrated limited cross-ancestry performance of PRSs Kachuri et al. [2024] leading to the development of several multi-ancestry methods, including PRS-CSx [Ruan et al., 2022], SDPRX [Zhou et al., 2023], MUSSEL [Jin et al., 2024], JointPRS [Xu et al., 2025], and MIXPRS [Xu et al., 2026]. While we do not benchmark these methods here, we provide a strong infrastructure for the clean benchmarking of these methods internally. Because family graphs are constructed across ancestries, the evaluation cohorts for each ancestry are leakage-safe with respect to the discovery cohorts of all other ancestries. We leave the benchmarking of these and other multiancestry methods to future work.

The present benchmark has several limitations. First, EHR phenotypes contain diagnostic heterogeneity and trait-dependent observation bias. Second, the discovery sample is necessarily smaller than the full AoU cohort, which limits power for rare diseases and makes internal-discovery-only comparisons sensitive to case count. Third, ancestry groups differ in sample size and disease prevalence, so not every internal-discovery-only task will be equally powered across populations. Fourth, within-family estimates remain less precise than population estimates for rare traits due to low sample count and case count. Finally, benchmark results depend on the frozen phenotype definitions, prevalence assumptions, and evaluation metrics; these must be versioned rather than changed opportunistically as models evolve.

These constraints are features of the scientific problem rather than reasons to avoid benchmarking. By making the constraints explicit, the resource can distinguish gains due to statistical methodology from gains due to additional training data, altered phenotyping, or inadvertent leakage. The intended endpoint is a benchmark in which a new genomic method can be evaluated along a sequence of increasingly stringent questions: does it learn efficiently from internal-discovery-only data, does it improve over externally trained PRSs, does it transport across ancestry, does it retain signal within families, and does the gain persist in a locked reserve that was never used during development?

## Data availability

Individual-level AoU data are available to authorized researchers through the All of Us Researcher Workbench subject to program access requirements. All external GWAS summary statistics used in the baseline models are publicly available and downloadable from the original studies and consortia.

## Acknowledgments

We gratefully acknowledge the participants of the All of Us Research Program and the investigators and participants of the GWAS studies whose summary statistics contributed to this work, including but not limited to UKBB, MVP, FinnGen, and trait-specific consortia including but not limited to the European Alzheimer and Dementia Biobank [Bellenguez et al., 2022, EADB et al., 2026], Breast Cancer Association Consortium [Zhang et al., 2020], International IBD Genetics Consortium [De Lange et al., 2017], and Type 2 Diabetes Global Genomics Initiative [Suzuki et al., 2024]. All trait-specific consortia are provided in Table B5.

## Appendix

### A Evaluation Settings

#### A.1 Conversion between observed-scale and liability-scale variance

For a binary trait with population or lifetime prevalence *K*, sample case fraction *P*, observed-scale incremental variance explained *R^2^_O_*, threshold

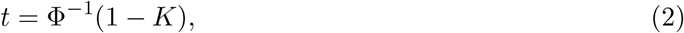

and standard-normal density *z* = *ϕ*(*t*), we used the Lee et al. liability-threshold transformation [Lee et al., 2012]. Defining

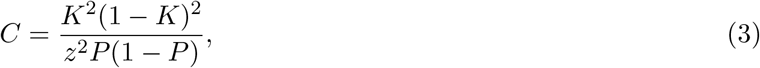

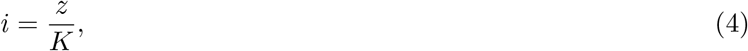

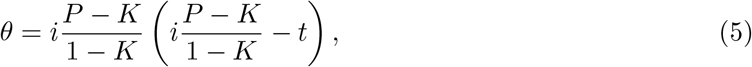

we compute

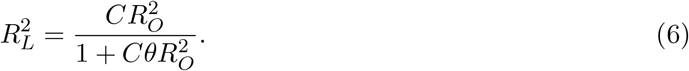

Trait-specific prevalence assumptions were frozen as part of the benchmark version. We matched the same lifetime risk for EUR-ancestry as in Moore et al. [2025] for all traits. Lifetime risk estimates for other ancestries are provided in Table B6.

#### A.2 Within-family evaluation settings

Scores for probands and parents were computed by SNIPAR [Young et al., 2022] with identical weight vectors and frozen normalization across family members. Parental genotyping in the holdout is sparse. A majority of modeled probands contain neither parent genotyped, with maternal and paternal scores collinear by construction, while few contain two genotyped parents (Table C4). Modeled rows required complete phenotype, proband score, age, and sex, and at least two modeled members per family. Traits used a probit GLM with family-cluster-robust standard errors (cluster unit: family ID). Estimates were flagged non-interpretable when the *β*_pop_ interval included zero. Estimates made from 20 or fewer cases are suppressed. Parental-adjustment (Young) estimates restricted to the ≥ 1-genotyped-parent stratum and per-stratum sibship-contrast estimates are reported in Table C4.

### B Supplementary tables

#### B.1 Phenotype definitions

**Table B1:** Phenotype definitions and control criteria across traits. Concept IDs are OMOP concepts (SNOMED vocabulary unless noted) as resolved under the All of Us C2025Q4R6 curated dataset. “Root + descendants” includes all active standard descendants of the listed concept in the extraction CDR. Case concepts and control exclusions are designed to avoid phenotype misclassification.

| Trait | Case concept(s) | Control exclusions |
| --- | --- | --- |
| Alzheimer's disease | 378419 (Alzheimer's disease), including descendants | Any dementia, MCI, or major-neurocognitive-disorder condition based on name pattern; controls age $\geq 65$ |
| Basal cell carcinoma | 109 SNOMED concepts under root 4112752 (Basal cell carcinoma of skin) (Table D1) plus 6 BCC-specific procedures (Table D2) | None |
| Breast cancer | 256 primary-malignancy concepts under 4157332 (Malignant neoplasm of female breast) (Table D3) | Secondary or metastatic breast evidence (11 concepts; Table D4). female sex |
| Glaucoma | 435262 (Primary open angle glaucoma) and 441561 (Low tension glaucoma), including descendants | Roots 437541, 3663197, and 381290 (secondary, neovascular, or borderline glaucoma); controls age $\geq 40$ with eye-exam (4263508, 4302595) |
| Gout | 440674 (Gout), including descendants | None |
| Hypertension | 16-concept essential-hypertension set (Table D5) | 115 blocking concepts (Table D6); controls age $\geq 50$ |
| Inflammatory bowel disease | 62-concept union of Crohn's disease, ulcerative colitis, and indeterminate IBD (Table D7) | 11 nonspecific-colitis exclusion concepts (Table D8) |
| Melanoma | 222 invasive cutaneous melanoma concepts (Table D9) | Any reviewed melanoma evidence; controls with age $\geq q10$ case-age floor fit on discovery |
| Multiple sclerosis | 15 explicit concepts (Table D10) | Roots 37116351 (Factor VIII deficiency syndrome) and 44811462 (Uhthoff Phenomenon) |
| Osteoporosis | 80502 (Osteoporosis), including descendants | None |
| Prostate cancer | 93 direct or advanced prostate-malignancy concepts (Table D11) | 102 indeterminate concepts block controls (Table D12); male sex |
| Psoriasis | 140168 (Psoriasis), including descendants | None |
| Testicular cancer | 65 TGCT or broad-malignancy diagnosis concepts (Table D13) | Male sex; age $\geq 18$ |
| Type 2 diabetes | 201826 (Type 2 diabetes mellitus), including descendants | None |

#### B.2 Case count for discovery phenotypes

**Table B2:**
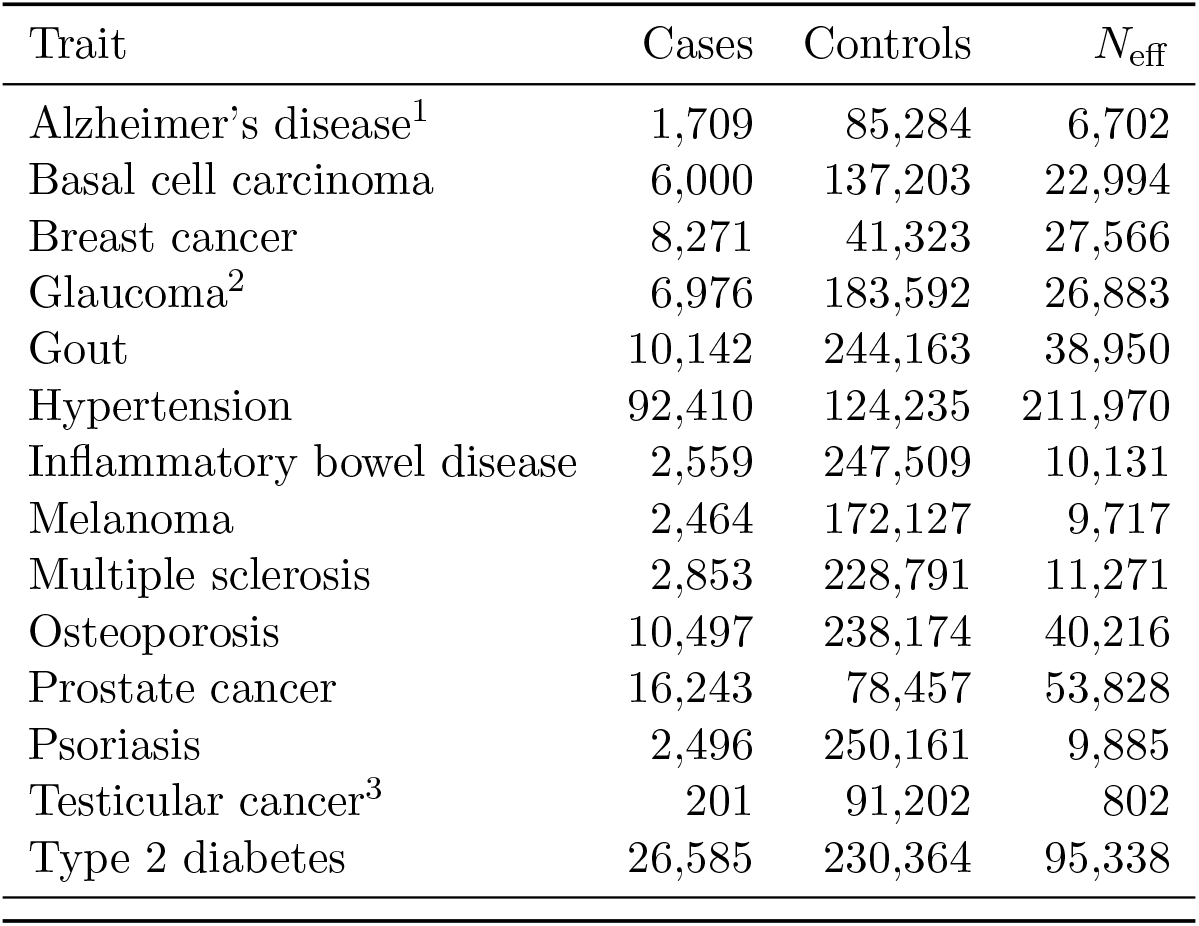
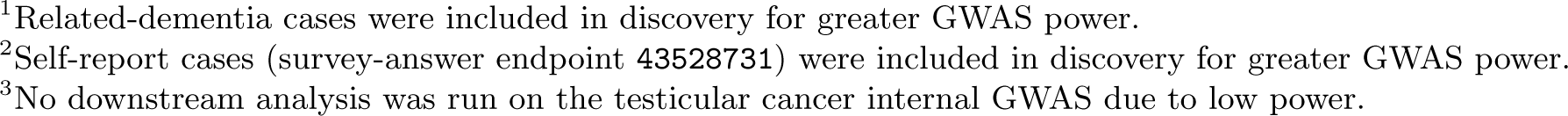
Phenotype sample sizes and effective sample sizes for discovery cohort internal GWAS.

#### B.3 Case count for external-permitted scores

**Table B3:**
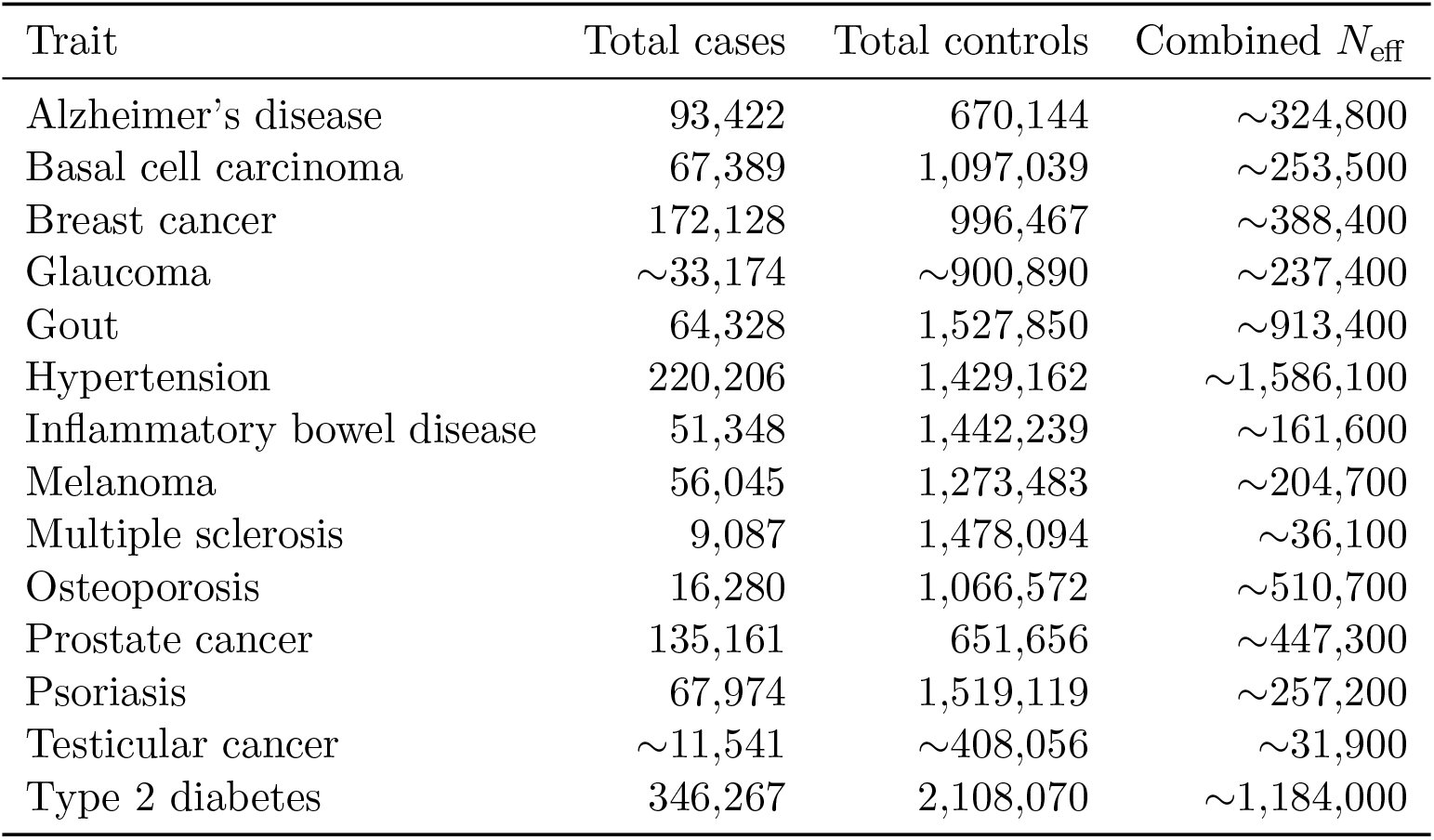
Summary of combined meta-analysis sample sizes used for polygenic risk score construction for the external-permitted track.

#### B.4 Cross-ancestry transport case counts

**Table B4:** Locked-test sample sizes and case counts for the cross-ancestry score-only AUC analysis. Entries are reported as total *n* / number of cases. Case counts of 20 or under are suppressed as per the AoU data privacy agreement.

| Trait | EUR | AFR | AMR | EAS | MID | SAS |
| --- | --- | --- | --- | --- | --- | --- |
| Alzheimer’s disease | 23,404 / 144 | 4,072 / $\leq 20$ | 2,635 / 26 | 470 / $\leq 20$ | 90 / $\leq 20$ | 130 / $\leq 20$ |
| Basal cell carcinoma | 34,552 / 2,057 | 9,193 / $\leq 20$ | 6,237 / 50 | 990 / $\leq 20$ | 151 / $\leq 20$ | 287 / $\leq 20$ |
| Breast cancer | 29,754 / 1,857 | 8,553 / 324 | 8,891 / 290 | 1,479 / 56 | 176 / $\leq 20$ | 386 / $\leq 20$ |
| Glaucoma | 5,575 / 850 | 940 / 377 | 900 / 155 | 154 / 38 | 28 / $\leq 20$ | 60 / $\leq 20$ |
| Gout | 54,954 / 1,559 | 15,697 / 473 | 15,267 / 193 | 2,637 / 87 | 372 / $\leq 20$ | 898 / $\leq 20$ |
| Hypertension | 36,626 / 20,649 | 10,413 / 6,805 | 7,559 / 4,546 | 1,142 / 559 | 180 / 94 | 328 / 175 |
| Inflammatory bowel disease | 54,639 / 746 | 15,806 / 84 | 15,225 / 86 | 2,638 / $\leq 20$ | 373 / $\leq 20$ | 901 / $\leq 20$ |
| Melanoma | 40,106 / 503 | 11,374 / $\leq 20$ | 8,635 / $\leq 20$ | 1,381 / $\leq 20$ | 207 / $\leq 20$ | 421 / $\leq 20$ |
| Multiple sclerosis | 50,839 / 582 | 14,791 / 117 | 13,726 / 81 | 2,291 / $\leq 20$ | 344 / $\leq 20$ | 770 / $\leq 20$ |
| Osteoporosis | 54,361 / 3,073 | 15,767 / 268 | 15,161 / 325 | 2,606 / 71 | 373 / $\leq 20$ | 889 / $\leq 20$ |
| Prostate cancer | 21,365 / 4,552 | 6,091 / 615 | 4,882 / 485 | 817 / 84 | 169 / 21 | 383 / 25 |
| Psoriasis | 55,338 / 731 | 15,889 / 55 | 15,326 / 112 | 2,639 / 23 | 376 / $\leq 20$ | 900 / $\leq 20$ |
| Testicular cancer | 20,511 / 56 | 5,968 / $\leq 20$ | 4,714 / $\leq 20$ | 793 / $\leq 20$ | 161 / $\leq 20$ | 378 / $\leq 20$ |
| Type 2 diabetes | 56,423 / 4,999 | 15,952 / 2,045 | 15,602 / 1,855 | 2,768 / 193 | 387 / 39 | 956 / 87 |

#### B.5 Provenance for external consortia GWAS

**Table B5:** External non-biobank GWAS analyses used for polygenic score construction. Where available, GWAS summmary statistics from UK Biobank, FinnGen, and MVP are used as well. These are not documented in this provenance table for brevity.

| Trait | External GWAS Source |
| --- | --- |
| Alzheimer’s disease | EADB et al. [2026] |
| Basal cell carcinoma | Seviiri et al. [2022] |
| Breast cancer | Zhang et al. [2020] |
| Glaucoma | Gharahkhani et al. [2021], Han et al. [2021] |
| Gout | Cho et al. [2024] |
| Hypertension | Keaton et al. [2024] |
| Inflammatory bowel disease | De Lange et al. [2017] |
| Prostate cancer | Wang et al. [2023] |
| Psoriasis | Dand et al. [2025] |
| Testicular cancer | Wang et al. [2017] |
| Type 2 diabetes | Suzuki et al. [2024] |

#### B.6 Lifetime risk estimates for non-EUR ancestry

**Table B6:** Cited lifetime risk (prevalence) *K* used for multi-ancestry liability-scale transport, by trait and ancestry proxy.

| Trait | Ancestry | $K$ (%) | Source |
| --- | --- | --- | --- |
| Alzheimer’s disease | EAS | 31.0 | Yoshida et al. [2020] |
| Breast cancer | AFR | 11.6 | National Cancer Institute [2021] |
|  | AMR | 10.8 | National Cancer Institute [2021] |
|  | EAS | 4.2 | Liao et al. [2025] |
| Glaucoma | AFR | 11.5 | Friedman et al. [2004] |
|  | AMR | 9.0 | Friedman et al. [2004] |
| Gout | AFR | 5.9 | Yokose et al. [2023] |
|  | AMR | 3.9 | Yokose et al. [2023] |
|  | EAS | 6.3 | Kuo et al. [2015] |
| Hypertension | AFR | 76 | Chen et al. [2019] |
|  | AMR | 92 | Carson et al. [2011] |
|  | SAS | 79 | Kaur et al. [2022] |
|  | EAS | 84 | Carson et al. [2011] |
| Inflammatory bowel disease | AFR | 0.50 | Lewis et al. [2023] |
|  | AMR | 0.46 | Lewis et al. [2023] |
| Multiple sclerosis | AFR | 0.29 | Hittle et al. [2023] |
|  | AMR | 0.16 | Hittle et al. [2023] |
|  | MID | 0.08 | Alroughani et al. [2014] |
| Prostate cancer | AFR | 16.7 | National Cancer Institute [2021] |
|  | AMR | 11.1 | National Cancer Institute [2021] |
|  | EAS | 8.7 | Miller et al. [2006] |
| Psoriasis | AFR | 1.5 | Armstrong et al. [2021] |
|  | AMR | 1.9 | Armstrong et al. [2021] |
|  | EAS | 0.38 | Choon et al. [2022] |
| Type 2 diabetes | AFR | 27.5 | Narayan et al. [2003] |
|  | AMR | 48.9 | Narayan et al. [2003] |
|  | SAS | 25.3 | Narayan et al. [2003] |
|  | EAS | 25.7 | Narayan et al. [2003] |
|  | MID | 59.6 | Ramezankhani et al. [2022] |

### C Supplementary results

#### C.1 Liability-scale variance explained for external-permitted PRSs

**Table C1:** Liability-scale variance explained for our meta-analyses across 14 binary trait endpoints.

| Trait | Liability $R^2$ [95% CI] |
| --- | --- |
| Alzheimer’s disease | 24.91 [13.66, 38.41] |
| Basal cell carcinoma | 13.76 [12.07, 15.52] |
| Breast cancer | 12.14 [10.29, 14.00] |
| Glaucoma | 16.28 [13.75, 18.93] |
| Gout | 13.14 [11.49, 14.84] |
| Hypertension | 11.83 [11.04, 12.60] |
| Inflammatory bowel disease | 14.63 [12.45, 16.83] |
| Melanoma | 7.69 [5.76, 10.05] |
| Multiple sclerosis | 11.02 [9.13, 13.07] |
| Osteoporosis | 5.73 [4.83, 6.73] |
| Prostate cancer | 17.32 [15.52, 19.15] |
| Psoriasis | 9.28 [7.33, 11.35] |
| Testicular cancer | 12.86 [7.69, 19.49] |
| Type 2 diabetes | 18.05 [16.82, 19.23] |

#### C.2 Liability-scale variance explained for internal-discovery-only PRSs

**Table C2:** Performance of internal-discovery-only polygenic scores across traits. For each trait, we report the total number of individuals in the test sample, the population prevalence matching [Moore et al., 2025], the corresponding *R*^2^, and the predictive score-only AUC. No results were computed for testicular cancer due to the lack of a meaningful internal GWAS. Predictive AUC was only computed on the covariate-complete subset of the test set.

| Trait | $n_{\text{test}}$ | $K$ | Internal-discovery-only<br>$R_L^2$ % [95% CI] | Predictive AUC |
| --- | --- | --- | --- | --- |
| Alzheimer’s disease | 23,149 | 0.111 | **14.51** [7.01, 24.19] | 0.650 |
| Basal cell carcinoma | 34,216 | 0.195 | **7.52** [6.17, 8.96] | 0.636 |
| Breast cancer | 29,754 | 0.1387 | **1.15** [0.67, 1.73] | 0.567 |
| Glaucoma | 5,522 | 0.0702 | **2.68** [1.72, 3.89] | 0.599 |
| Gout | 54,511 | 0.0705 | **5.88** [4.69, 7.14] | 0.632 |
| Hypertension | 36,279 | 0.4685 | **5.75** [5.18, 6.37] | 0.602 |
| Inflammatory bowel disease | 54,196 | 0.0177 | **3.62** [2.60, 4.79] | 0.634 |
| Melanoma | 39,742 | 0.02975 | **5.17** [3.45, 7.05] | 0.635 |
| Multiple sclerosis | 50,419 | 0.00365 | **6.22** [4.80, 7.79] | 0.680 |
| Osteoporosis | 53,920 | 0.34 | **1.74** [1.20, 2.35] | 0.570 |
| Prostate cancer | 21,365 | 0.1257 | **2.79** [2.26, 3.38] | 0.587 |
| Psoriasis | 54,886 | 0.029 | **2.78** [1.79, 4.07] | 0.605 |
| Type 2 diabetes | 55,971 | 0.199 | **7.13** [6.30, 7.98] | 0.624 |
| Testicular cancer | — | — | *No internal GWAS analyses* | — |

#### C.3 Cross-ancestry liability-scale variance explained for external-permitted PRS

**Figure 4:**
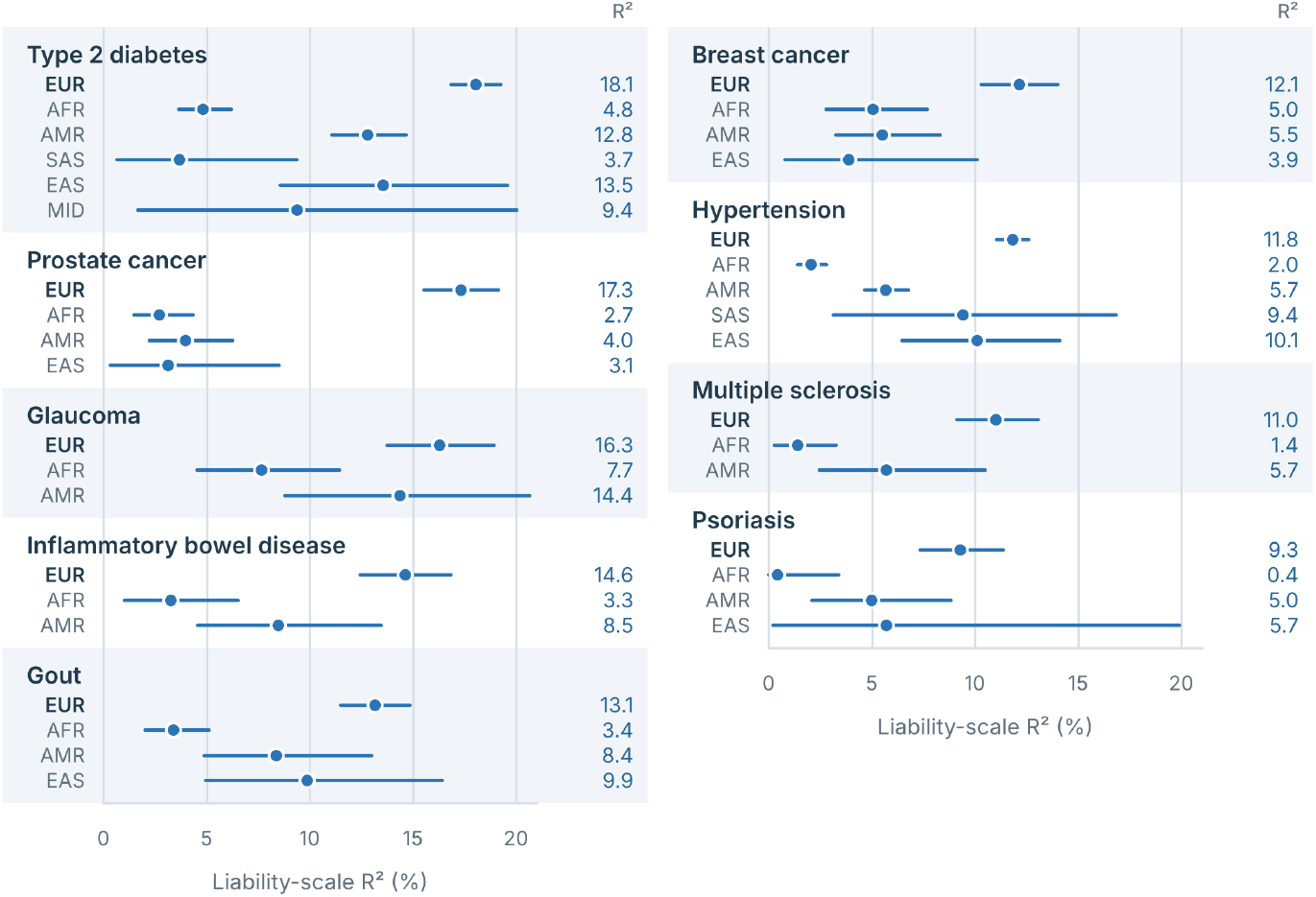
Cross-ancestry liability-scale *R*^2^ results for 9 traits in EUR and non-EUR ancestry for external-permitted PRSs. Non-EUR lifetime risk estimates and sources are provided in Table B6.

#### C.4 Internal-discovery-only cross-ancestry transport

**Table C3:** Cross-ancestry score-only AUC for EUR-ancestry AoU internal-discovery-only PRSs. 95% confidence intervals were computed on 2000-replicate stratified bootstraps. Score-only AUC was computed on the entire test set, including individuals missing covariate information that were excluded from other evaluation settings. Estimates made from 20 or fewer cases are suppressed. No internal testicular cancer PRS was trained due to statistical non-interpretability.

| Trait | EUR | AFR | AMR | EAS | MID | SAS |
| --- | --- | --- | --- | --- | --- | --- |
| Alzheimer's disease | 0.648 [0.600, 0.699] | — | 0.705 [0.620, 0.785] | — | — | — |
| Basal cell carcinoma | 0.635 [0.624, 0.648] | — | 0.667 [0.583, 0.743] | — | — | — |
| Breast cancer | 0.567 [0.554, 0.580] | 0.490 [0.456, 0.522] | 0.527 [0.494, 0.559] | 0.462 [0.382, 0.545] | — | — |
| Glaucoma | 0.600 [0.579, 0.621] | 0.581 [0.543, 0.617] | 0.603 [0.553, 0.652] | 0.535 [0.438, 0.636] | — | — |
| Gout | 0.632 [0.618, 0.646] | 0.563 [0.538, 0.589] | 0.646 [0.607, 0.685] | 0.662 [0.610, 0.717] | — | — |
| Hypertension | 0.601 [0.595, 0.607] | 0.534 [0.522, 0.545] | 0.601 [0.589, 0.615] | 0.623 [0.590, 0.655] | 0.632 [0.547, 0.710] | 0.588 [0.529, 0.650] |
| Inflammatory bowel disease | 0.635 [0.614, 0.654] | 0.563 [0.506, 0.618] | 0.623 [0.562, 0.687] | — | — | — |
| Melanoma | 0.635 [0.610, 0.660] | — | — | — | — | — |
| Multiple sclerosis | 0.679 [0.656, 0.702] | 0.560 [0.509, 0.611] | 0.610 [0.544, 0.675] | — | — | — |
| Osteoporosis | 0.570 [0.559, 0.581] | 0.543 [0.509, 0.578] | 0.572 [0.541, 0.602] | 0.556 [0.487, 0.623] | — | — |
| Prostate cancer | 0.587 [0.578, 0.596] | 0.532 [0.508, 0.555] | 0.547 [0.519, 0.573] | 0.548 [0.482, 0.615] | 0.603 [0.448, 0.748] | 0.511 [0.380, 0.642] |
| Psoriasis | 0.604 [0.584, 0.625] | 0.489 [0.409, 0.570] | 0.560 [0.505, 0.617] | 0.664 [0.544, 0.778] | — | — |
| Type 2 diabetes | 0.624 [0.617, 0.632] | 0.545 [0.532, 0.558] | 0.592 [0.579, 0.606] | 0.627 [0.585, 0.668] | 0.570 [0.464, 0.666] | 0.554 [0.495, 0.612] |

#### C.5 Within-family evaluation for external-permitted PRSs

**Table C4:** Within-family effect estimates by parental-genotype stratum for 13 binary traits, with testicular cancer excluded due to low case count. Entries give the point estimate over the 95% bootstrap CI. *R*^sib^_attenuation_ is the sibship-contrast ratio *β*_within_/*β*_pop_ (2,000 whole-family bootstrap replicates); *R*^par^_attenuation_ is the Young parental-adjustment ratio restricted to probands with ≥ 1 genotyped parent (also 2,000 replicates). The first two estimate columns are *R*^sib^_attenuation_; the last is *R*^par^_attenuation_. Alzheimer’s disease all-sibship ratio was statistically non-interpretable. *n*_mod_ refers to the number of modeled probands. Parental-genotype composition is given as coarsened percentages of modeled probands; cells below the disclosure threshold are suppressed.

| Trait | $n_{\text{mod}}$ | Parents genotyped (%) | | | sibship contrast | | parental adj. |
| --- | --- | --- | --- | --- | --- | --- | --- |
| | | both | one | neither | all sibships | no parent | $\geq 1$ parent |
| Alzheimer’s disease | 3009 | <1 | 3 | 97 | 0.60<br>[−6.30, 5.38] | — | — |
| Basal cell carcinoma | 6003 | 1 | 9 | 90 | 1.07<br>[0.78, 1.39] | 1.01<br>[0.69, 1.35] | — |
| Breast cancer | 7120 | 3 | 29 | 68 | 1.08<br>[0.78, 1.39] | 1.01<br>[0.54, 1.54] | — |
| Glaucoma | 9015 | 2 | 19 | 79 | 0.96<br>[0.67, 1.29] | 0.85<br>[0.36, 1.30] | — |
| Gout | 15345 | 3 | 28 | 69 | 1.09<br>[0.86, 1.33] | 1.02<br>[0.72, 1.34] | — |
| Hypertension | 13715 | 3 | 28 | 69 | 0.91<br>[0.80, 1.01] | 1.00<br>[0.85, 1.16] | 0.96<br>[0.77, 1.16] |
| Inflammatory bowel disease | 14786 | 3 | 28 | 68 | 1.13<br>[0.86, 1.41] | 1.00<br>[0.61, 1.43] | — |
| Melanoma | 8097 | 2 | 15 | 83 | 1.24<br>[0.79, 1.84] | 1.18<br>[0.67, 1.79] | — |
| Multiple sclerosis | 13598 | 3 | 28 | 69 | 0.90<br>[0.56, 1.25] | 1.16<br>[0.75, 1.72] | — |
| Osteoporosis | 14552 | 3 | 29 | 68 | 0.97<br>[0.73, 1.20] | 1.00<br>[0.68, 1.32] | — |
| Prostate cancer | 1279 | 3 | 27 | 70 | 1.11<br>[0.72, 1.60] | 0.83<br>[0.38, 1.55] | — |
| Psoriasis | 15113 | 3 | 28 | 69 | 0.94<br>[0.62, 1.25] | 0.64<br>[0.02, 1.13] | — |
| Type 2 diabetes | 15488 | 3 | 28 | 68 | 0.86<br>[0.76, 0.97] | 0.86<br>[0.70, 1.03] | 1.02<br>[0.86, 1.19] |

### D Specific trait concept roots

#### D.1 Basal cell carcinoma

**Table D1:**
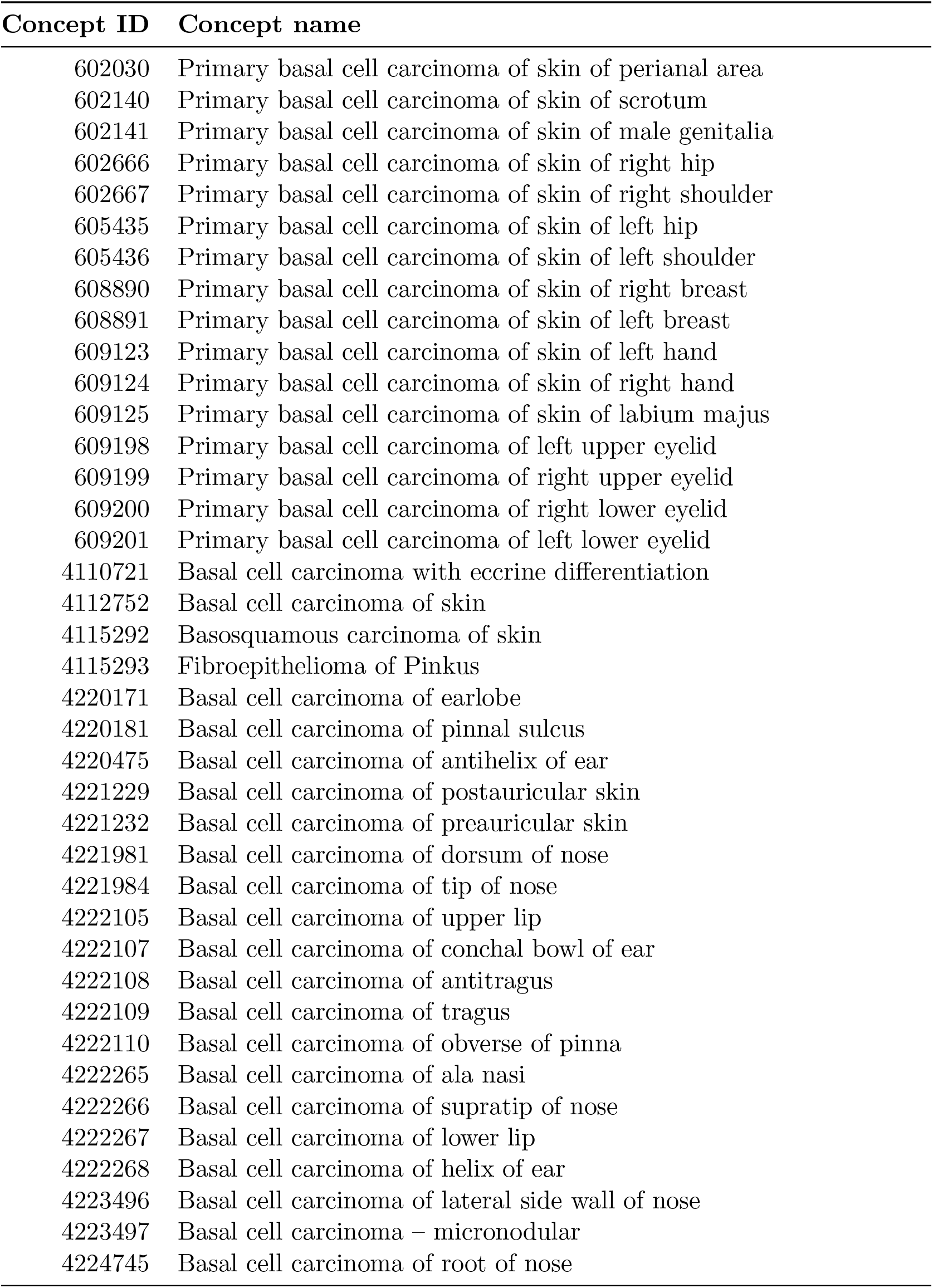
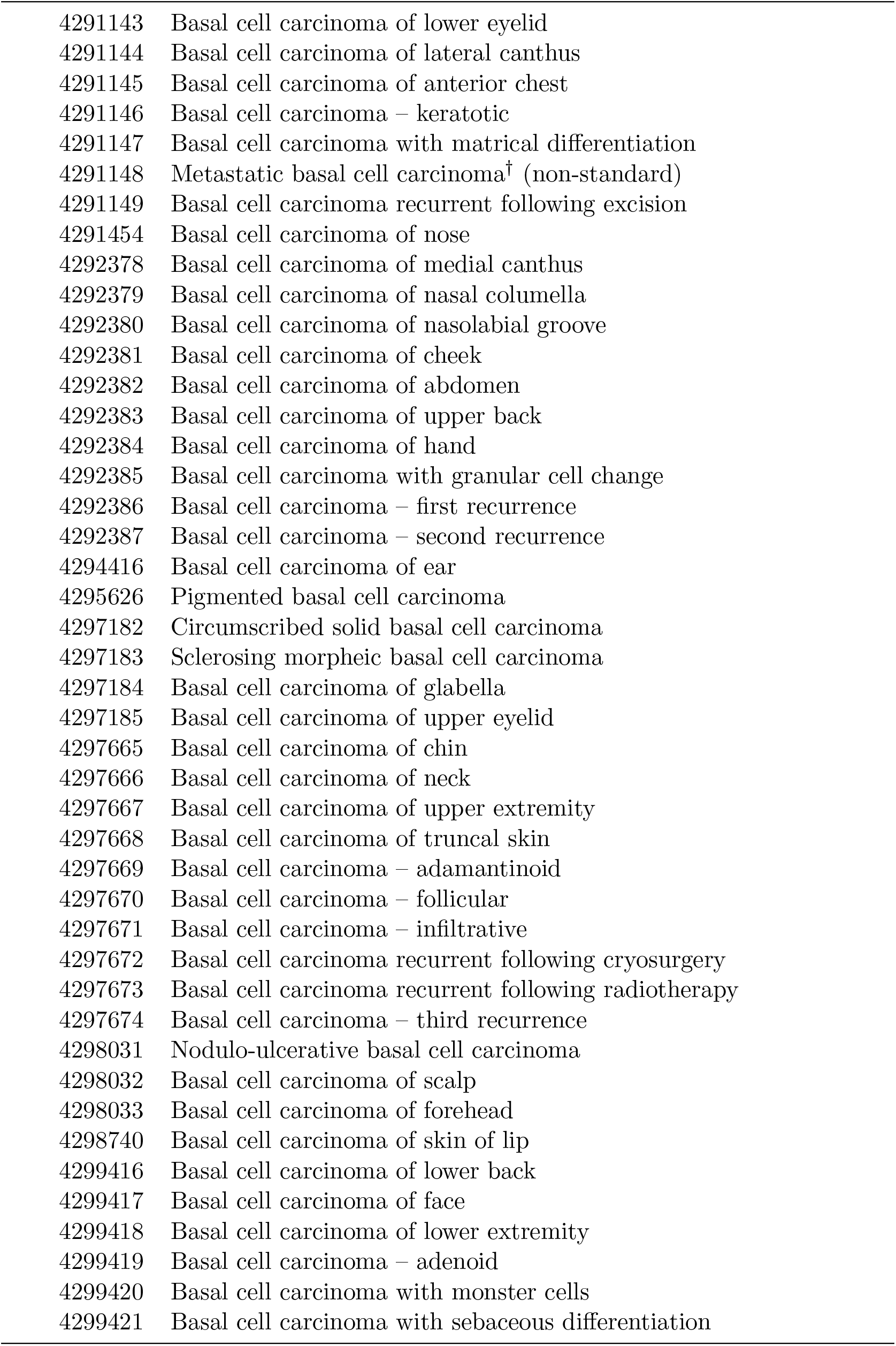
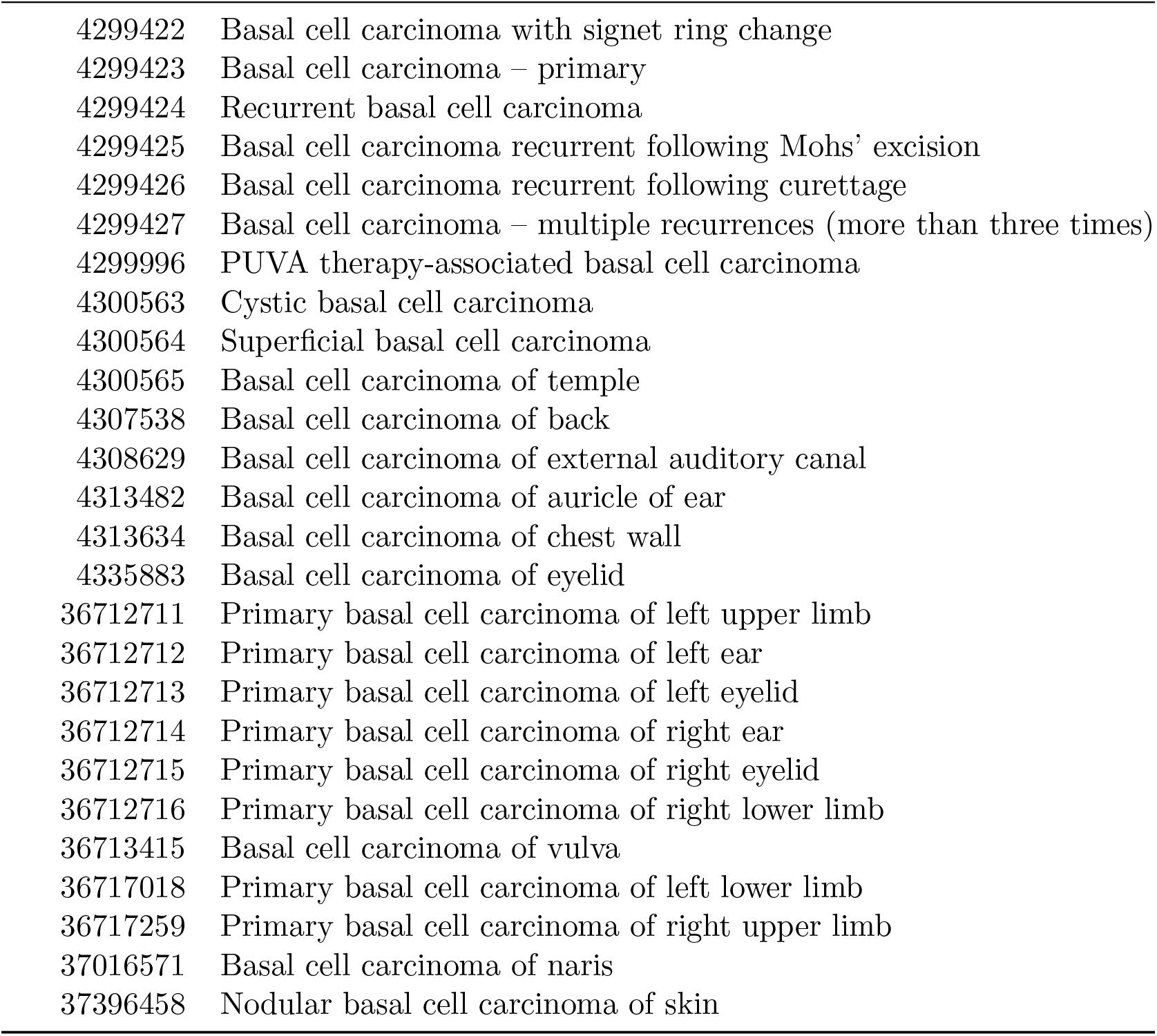
Basal cell carcinoma case condition concepts.

| Concept ID | Concept name |
| --- | --- |
| 602030 | Primary basal cell carcinoma of skin of perianal area |
| 602140 | Primary basal cell carcinoma of skin of scrotum |
| 602141 | Primary basal cell carcinoma of skin of male genitalia |
| 602666 | Primary basal cell carcinoma of skin of right hip |
| 602667 | Primary basal cell carcinoma of skin of right shoulder |
| 605435 | Primary basal cell carcinoma of skin of left hip |
| 605436 | Primary basal cell carcinoma of skin of left shoulder |
| 608890 | Primary basal cell carcinoma of skin of right breast |
| 608891 | Primary basal cell carcinoma of skin of left breast |
| 609123 | Primary basal cell carcinoma of skin of left hand |
| 609124 | Primary basal cell carcinoma of skin of right hand |
| 609125 | Primary basal cell carcinoma of skin of labium majus |
| 609198 | Primary basal cell carcinoma of left upper eyelid |
| 609199 | Primary basal cell carcinoma of right upper eyelid |
| 609200 | Primary basal cell carcinoma of right lower eyelid |
| 609201 | Primary basal cell carcinoma of left lower eyelid |
| 4110721 | Basal cell carcinoma with eccrine differentiation |
| 4112752 | Basal cell carcinoma of skin |
| 4115292 | Basosquamous carcinoma of skin |
| 4115293 | Fibroepithelioma of Pinkus |
| 4220171 | Basal cell carcinoma of earlobe |
| 4220181 | Basal cell carcinoma of pinna sulcus |
| 4220475 | Basal cell carcinoma of antihelix of ear |
| 4221229 | Basal cell carcinoma of postauricular skin |
| 4221232 | Basal cell carcinoma of preauricular skin |
| 4221981 | Basal cell carcinoma of dorsum of nose |
| 4221984 | Basal cell carcinoma of tip of nose |
| 4222105 | Basal cell carcinoma of upper lip |
| 4222107 | Basal cell carcinoma of conchal bowl of ear |
| 4222108 | Basal cell carcinoma of antitragus |
| 4222109 | Basal cell carcinoma of tragus |
| 4222110 | Basal cell carcinoma of obverse of pinna |
| 4222265 | Basal cell carcinoma of ala nasi |
| 4222266 | Basal cell carcinoma of supratip of nose |
| 4222267 | Basal cell carcinoma of lower lip |
| 4222268 | Basal cell carcinoma of helix of ear |
| 4223496 | Basal cell carcinoma of lateral side wall of nose |
| 4223497 | Basal cell carcinoma – micronodular |
| 4224745 | Basal cell carcinoma of root of nose |

Table D1 – continued from previous page
| Concept ID | Concept name |
| --- | --- |
| 4291143 | Basal cell carcinoma of lower eyelid |
| 4291144 | Basal cell carcinoma of lateral canthus |
| 4291145 | Basal cell carcinoma of anterior chest |
| 4291146 | Basal cell carcinoma – keratotic |
| 4291147 | Basal cell carcinoma with matrical differentiation |
| 4291148 | Metastatic basal cell carcinoma<sup>†</sup> (non-standard) |
| 4291149 | Basal cell carcinoma recurrent following excision |
| 4291454 | Basal cell carcinoma of nose |
| 4292378 | Basal cell carcinoma of medial canthus |
| 4292379 | Basal cell carcinoma of nasal columella |
| 4292380 | Basal cell carcinoma of nasolabial groove |
| 4292381 | Basal cell carcinoma of cheek |
| 4292382 | Basal cell carcinoma of abdomen |
| 4292383 | Basal cell carcinoma of upper back |
| 4292384 | Basal cell carcinoma of hand |
| 4292385 | Basal cell carcinoma with granular cell change |
| 4292386 | Basal cell carcinoma – first recurrence |
| 4292387 | Basal cell carcinoma – second recurrence |
| 4294416 | Basal cell carcinoma of ear |
| 4295626 | Pigmented basal cell carcinoma |
| 4297182 | Circumscribed solid basal cell carcinoma |
| 4297183 | Sclerosing morpheic basal cell carcinoma |
| 4297184 | Basal cell carcinoma of glabella |
| 4297185 | Basal cell carcinoma of upper eyelid |
| 4297665 | Basal cell carcinoma of chin |
| 4297666 | Basal cell carcinoma of neck |
| 4297667 | Basal cell carcinoma of upper extremity |
| 4297668 | Basal cell carcinoma of truncal skin |
| 4297669 | Basal cell carcinoma – adamantinoid |
| 4297670 | Basal cell carcinoma – follicular |
| 4297671 | Basal cell carcinoma – infiltrative |
| 4297672 | Basal cell carcinoma recurrent following cryosurgery |
| 4297673 | Basal cell carcinoma recurrent following radiotherapy |
| 4297674 | Basal cell carcinoma – third recurrence |
| 4298031 | Nodulo-ulcerative basal cell carcinoma |
| 4298032 | Basal cell carcinoma of scalp |
| 4298033 | Basal cell carcinoma of forehead |
| 4298740 | Basal cell carcinoma of skin of lip |
| 4299416 | Basal cell carcinoma of lower back |
| 4299417 | Basal cell carcinoma of face |
| 4299418 | Basal cell carcinoma of lower extremity |
| 4299419 | Basal cell carcinoma – adenoid |
| 4299420 | Basal cell carcinoma with monster cells |
| 4299421 | Basal cell carcinoma with sebaceous differentiation |

Table D1 – continued from previous page
| Concept ID | Concept name |
| --- | --- |
| 4299422 | Basal cell carcinoma with signet ring change |
| 4299423 | Basal cell carcinoma – primary |
| 4299424 | Recurrent basal cell carcinoma |
| 4299425 | Basal cell carcinoma recurrent following Mohs’ excision |
| 4299426 | Basal cell carcinoma recurrent following curettage |
| 4299427 | Basal cell carcinoma – multiple recurrences (more than three times) |
| 4299996 | PUVA therapy-associated basal cell carcinoma |
| 4300563 | Cystic basal cell carcinoma |
| 4300564 | Superficial basal cell carcinoma |
| 4300565 | Basal cell carcinoma of temple |
| 4307538 | Basal cell carcinoma of back |
| 4308629 | Basal cell carcinoma of external auditory canal |
| 4313482 | Basal cell carcinoma of auricle of ear |
| 4313634 | Basal cell carcinoma of chest wall |
| 4335883 | Basal cell carcinoma of eyelid |
| 36712711 | Primary basal cell carcinoma of left upper limb |
| 36712712 | Primary basal cell carcinoma of left ear |
| 36712713 | Primary basal cell carcinoma of left eyelid |
| 36712714 | Primary basal cell carcinoma of right ear |
| 36712715 | Primary basal cell carcinoma of right eyelid |
| 36712716 | Primary basal cell carcinoma of right lower limb |
| 36713415 | Basal cell carcinoma of vulva |
| 36717018 | Primary basal cell carcinoma of left lower limb |
| 36717259 | Primary basal cell carcinoma of right upper limb |
| 37016571 | Basal cell carcinoma of naris |
| 37396458 | Nodular basal cell carcinoma of skin |

**Table D2:**
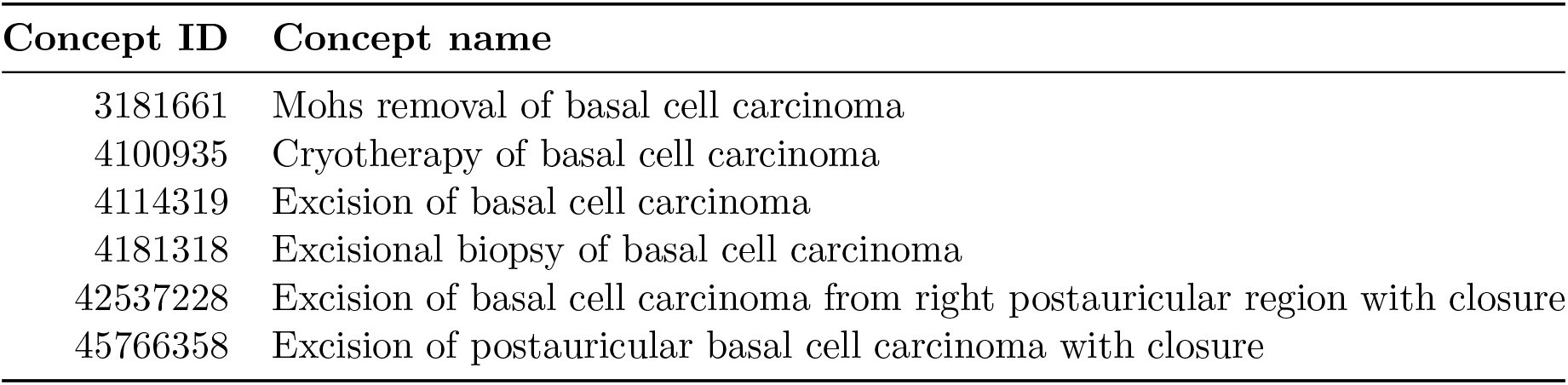
Basal cell carcinoma-specific procedure concepts.

#### D.2 Breast cancer

**Table D3:** 256 Primary breast malignancy case concepts.

| Concept ID | Concept name |
| --- | --- |
| 36402993 | Paget disease in situ and intraductal carcinoma (C50._) of nipple |
| 36403075 | Paget disease, mammary in situ (C50._) of nipple |
| 759932 | Infiltrating duct carcinoma of left female breast |
| 759933 | Infiltrating duct carcinoma of right female breast |
| 761170 | Infiltrating duct carcinoma of bilateral female breasts |
| 36517460 | Mucin-producing adenocarcinoma of nipple |
| 36517509 | Malignant tumor, clear cell type of nipple |
| 36517515 | Mucinous cystadenocarcinoma, NOS, of nipple |
| 36517806 | Hemangiosarcoma of nipple |
| 36517996 | Basal cell adenocarcinoma of nipple |
| 36518002 | Duct carcinoma, desmoplastic type of nipple |
| 36518332 | Infiltrating ductular carcinoma of nipple |
| 36518576 | Phyllodes tumor, malignant of nipple |
| 36518802 | Signet ring cell carcinoma of nipple |
| 36520200 | Squamous cell carcinoma, NOS, of nipple |
| 36520783 | Small cell sarcoma of nipple |
| 36521490 | Langerhans cell sarcoma of nipple |
| 36521708 | B lymphoblastic leukemia/lymphoma, NOS, of nipple |
| 36521743 | Fibroblastic reticular cell tumor of nipple |
| 36522135 | Solid carcinoma, NOS, of nipple |
| 36522597 | Intraductal papillary adenocarcinoma with invasion of nipple |
| 36522605 | Medullary carcinoma with lymphoid stroma of nipple |
| 36522838 | Large cell carcinoma, NOS, of nipple |
| 36523238 | Hodgkin sarcoma of nipple |
| 36523456 | Dedifferentiated liposarcoma of nipple |
| 36523513 | B lymphoblastic leukemia/lymphoma with t(v;11q23); MLL rearranged of nipple |
| 36523586 | Sezary syndrome of nipple |
| 36523645 | Follicular lymphoma, grade 3 of nipple |
| 36523688 | Verrucous carcinoma, NOS, of nipple |
| 36523859 | Mesenchymoma, malignant of nipple |
| 36524088 | Leiomyosarcoma, NOS, of nipple |
| 36524161 | Malignant lymphoma, NOS, of nipple |
| 36525094 | Precursor T-cell lymphoblastic leukemia of nipple |
| 36525641 | Epithelioid leiomyosarcoma of nipple |
| 36525676 | Epithelioid sarcoma, NOS, of nipple |
| 36525707 | Myxofibrosarcoma of nipple |
| 36525748 | T-cell/histiocyte rich large B-cell lymphoma of nipple |
| 36526585 | Polygonal cell carcinoma of nipple |
| 36527177 | Round cell liposarcoma of nipple |
| 36527390 | Hodgkin lymphoma, lymphocyte depletion, diffuse fibrosis of nipple |
| 36527656 | Mast cell sarcoma of nipple |

Table D3 – continued from previous page
| Concept ID | Concept name |
| --- | --- |
| 36528520 | Fibrosarcoma, NOS, of nipple |
| 36528635 | Infiltrating lobular mixed with other types of carcinoma of nipple |
| 36528685 | Lipid-rich carcinoma of nipple |
| 36529601 | Mature T-cell lymphoma, NOS, of nipple |
| 36529700 | Systemic EBV positive T-cell lymphoproliferative disease of childhood of nipple |
| 36530281 | Intravascular large B-cell lymphoma of nipple |
| 36530356 | Neuroendocrine carcinoma, NOS, of nipple |
| 36530388 | Myxoid liposarcoma of nipple |
| 36530409 | Secretory carcinoma of nipple |
| 36530435 | Squamous cell carcinoma, keratinizing, NOS, of nipple |
| 36530498 | Glassy cell carcinoma of nipple |
| 36530708 | Myeloid or lymphoid neoplasm with PDGFRA rearrangement of nipple |
| 36531090 | Polymorphous adenocarcinoma of nipple |
| 36531354 | Carcinoma with osteoclast-like giant cells of nipple |
| 36531426 | Rhabdomyosarcoma, NOS, of nipple |
| 36531652 | Squamous cell carcinoma, small cell, nonkeratinizing of nipple |
| 36532344 | Hodgkin lymphoma, lymphocyte-rich of nipple |
| 36532444 | Stromal sarcoma, NOS, of nipple |
| 36532451 | Giant cell sarcoma of nipple |
| 36532695 | Giant cell and spindle cell carcinoma of nipple |
| 36532838 | Angioimmunoblastic T-cell lymphoma of nipple |
| 36533887 | Carcinosarcoma, NOS, of nipple |
| 36534084 | Acinar cell cystadenocarcinoma of nipple |
| 36534130 | Plasmacytoma, extramedullary of nipple |
| 36534279 | Pleomorphic carcinoma of nipple |
| 36536105 | Epithelial-myoepithelial carcinoma of nipple |
| 36536346 | Hodgkin lymphoma, nodular sclerosis, cellular phase of nipple |
| 36536630 | Mantle cell lymphoma of nipple |
| 36536968 | B lymphoblastic leukemia/lymphoma with t(9;22)(q34;q11.2); BCR-ABL1 of nipple |
| 36537556 | Hodgkin lymphoma, lymphocyte depletion, NOS, of nipple |
| 36538039 | Myoepithelial carcinoma of nipple |
| 36538045 | Solitary fibrous tumor, malignant of nipple |
| 36538093 | Small cell carcinoma, fusiform cell of nipple |
| 36538776 | Myosarcoma of nipple |
| 36538924 | Hodgkin lymphoma, NOS, of nipple |
| 36538949 | Adenoid cystic carcinoma of nipple |
| 36539400 | Malignant histiocytosis of nipple |
| 36539962 | Spindle cell sarcoma of nipple |
| 36540505 | Hodgkin lymphoma, nodular sclerosis, NOS, of nipple |
| 36540867 | Papillary squamous cell carcinoma of nipple |
| 36540965 | Hodgkin lymphoma, nodular lymphocyte predominant of nipple |
| 36541084 | Osteosarcoma, NOS, of nipple |
| 36541280 | Burkitt lymphoma, NOS, of nipple |
| 36541476 | Metaplastic carcinoma, NOS, of nipple |

Table D3 – continued from previous page
| Concept ID | Concept name |
| --- | --- |
| 36541792 | Adenocarcinoma with spindle cell metaplasia of nipple |
| 36541890 | Liposarcoma, well differentiated, NOS, of nipple |
| 36542159 | Interdigitating dendritic cell sarcoma of nipple |
| 36542372 | Giant cell carcinoma of nipple |
| 36542432 | Squamous cell carcinoma, large cell, nonkeratinizing, NOS, of nipple |
| 36542493 | Hodgkin lymphoma, nodular sclerosis, grade 2 of nipple |
| 36542809 | Malignant tumor, giant cell type of nipple |
| 36543218 | Hodgkin granuloma of nipple |
| 36543496 | B lymphoblastic leukemia/lymphoma with t(5;14)(q31;q32); IL3-IGH of nipple |
| 36543766 | Desmoplastic small round cell tumor of nipple |
| 36543781 | Diffuse large B-cell lymphoma, NOS, of nipple |
| 36543906 | Myeloid or lymphoid neoplasm with FGFR1 abnormalities of nipple |
| 36544024 | Epithelioma, malignant of nipple |
| 36544249 | Follicular lymphoma, grade 1 of nipple |
| 36544449 | B lymphoblastic leukemia/lymphoma with hypodiploidy (Hypodiploid ALL) of nipple |
| 36544796 | Malignant lymphoma, non-Hodgkin, NOS, of nipple |
| 36545441 | Embryonal sarcoma of nipple |
| 36545927 | Hodgkin lymphoma, mixed cellularity, NOS, of nipple |
| 36546031 | Superficial spreading adenocarcinoma of nipple |
| 36546372 | Undifferentiated sarcoma of nipple |
| 36546405 | Small cell carcinoma, NOS, of nipple |
| 36547138 | Malignant tumor, spindle cell type of nipple |
| 36547472 | Follicular lymphoma, NOS, of nipple |
| 36547912 | Carcinoma, undifferentiated, NOS, of nipple |
| 36548236 | Pleomorphic liposarcoma of nipple |
| 36548604 | Malignant mastocytosis of nipple |
| 36549874 | Hemangioendothelioma, malignant of nipple |
| 36550109 | Cystadenocarcinoma, NOS, of nipple |
| 36550313 | Composite Hodgkin and non-Hodgkin lymphoma of nipple |
| 36550519 | Squamous cell carcinoma, microinvasive of nipple |
| 36550807 | Spindle cell carcinoma, NOS, of nipple |
| 36551299 | Adenocarcinoma with mixed subtypes of nipple |
| 36551719 | Adenocarcinoma with neuroendocrine differentiation of nipple |
| 36551860 | T-cell large granular lymphocytic leukemia of nipple |
| 36552133 | Alveolar soft part sarcoma of nipple |
| 36552161 | Squamous cell carcinoma, adenoid of nipple |
| 36552167 | B lymphoblastic leukemia/lymphoma with t(1;19)(q23;p13.3); E2A-PBX1 (TCF3-PBX1) of nipple |
| 36552573 | Squamous cell carcinoma, spindle cell of nipple |
| 36553240 | Myeloproliferative neoplasm, unclassifiable of nipple |
| 36553345 | Langerhans cell histiocytosis, disseminated of nipple |
| 36553527 | Medullary carcinoma, NOS, of nipple |
| 36553810 | Adenocarcinoma with squamous metaplasia of nipple |
| 36553938 | Fibroblastic liposarcoma of nipple |

Table D3 – continued from previous page
| Concept ID | Concept name |
| --- | --- |
| 36553980 | Carcinoma, anaplastic, NOS, of nipple |
| 36554216 | Malignant lymphoma, large B-cell, diffuse, immunoblastic, NOS, of nipple |
| 36554300 | NK/T-cell lymphoma, nasal and nasal type of nipple |
| 36554533 | Apocrine adenocarcinoma of nipple |
| 36554750 | Mixed liposarcoma of nipple |
| 36554874 | Cystic hypersecretory carcinoma of nipple |
| 36555654 | Papillary adenocarcinoma, NOS, of nipple |
| 36555657 | Carcinoma simplex of nipple |
| 36556060 | Epithelioid hemangioendothelioma, NOS, of nipple |
| 36556222 | Mixed cell adenocarcinoma of nipple |
| 36556445 | Malignant tumor, small cell type of nipple |
| 36556821 | Encapsulated papillary carcinoma with invasion of nipple |
| 36557153 | Adenocarcinoma with cartilaginous and osseous metaplasia of nipple |
| 36557205 | Anaplastic large cell lymphoma, T-cell and Null-cell type of nipple |
| 36557226 | Sarcoma, NOS, of nipple |
| 36557230 | Pseudosarcomatous carcinoma of nipple |
| 36557277 | Squamous cell carcinoma with horn formation of nipple |
| 36557321 | Oxyphilic adenocarcinoma of nipple |
| 36557877 | Lymphoplasmacytic lymphoma of nipple |
| 36559780 | Granular cell tumor, malignant of nipple |
| 36559859 | Large cell carcinoma with rhabdoid phenotype of nipple |
| 36559944 | B lymphoblastic leukemia/lymphoma with hyperdiploidy of nipple |
| 36560055 | Sebaceous carcinoma of nipple |
| 36560424 | Histiocytic sarcoma of nipple |
| 36561380 | Infantile fibrosarcoma of nipple |
| 36561381 | Clear cell adenocarcinoma, NOS, of nipple |
| 36562037 | Carcinosarcoma, embryonal of nipple |
| 36562231 | Marginal zone B-cell lymphoma, NOS, of nipple |
| 36562484 | B-cell chronic lymphocytic leukemia/small lymphocytic lymphoma of nipple |
| 36562616 | Neoplasm, malignant of nipple |
| 36563045 | Tumor cells, malignant of nipple |
| 36563176 | Malignant lymphoma, mixed small and large cell, diffuse of nipple |
| 36563615 | Large cell neuroendocrine carcinoma of nipple |
| 36563848 | Granular cell carcinoma of nipple |
| 36563907 | Hodgkin lymphoma, lymphocyte depletion, reticular of nipple |
| 36564008 | Follicular lymphoma, grade 2 of nipple |
| 36564200 | Fascial fibrosarcoma of nipple |
| 36564604 | Glycogen-rich carcinoma of nipple |
| 36564713 | Myxoid leiomyosarcoma of nipple |
| 36565000 | Liposarcoma, NOS, of nipple |
| 36565551 | Precursor cell lymphoblastic lymphoma, NOS, of nipple |
| 36566123 | Plasmacytoma, NOS, of nipple |
| 36566267 | Acinar cell carcinoma of nipple |
| 36566310 | Trabecular adenocarcinoma of nipple |

Table D3 – continued from previous page
| Concept ID | Concept name |
| --- | --- |
| 36566588 | Follicular dendritic cell sarcoma of nipple |
| 36566773 | Adenocarcinoma with apocrine metaplasia of nipple |
| 36566941 | Alveolar adenocarcinoma of nipple |
| 36567217 | Adenocarcinoma in villous adenoma of nipple |
| 36567418 | Comedocarcinoma, NOS, of nipple |
| 36567537 | Adenosquamous carcinoma of nipple |
| 36567866 | B lymphoblastic leukemia/lymphoma with t(12;21)(p13;q22); TEL-AML1 (ETV6-RUNX1) of nipple |
| 36567991 | Hodgkin lymphoma, nodular sclerosis, grade 1 of nipple |
| 36568025 | Angiomyosarcoma of nipple |
| 36712719 | Infiltrating ductal carcinoma of upper inner quadrant of left female breast |
| 36712720 | Infiltrating ductal carcinoma of upper outer quadrant of left female breast |
| 36712721 | Infiltrating ductal carcinoma of central portion of right female breast |
| 36712722 | Infiltrating ductal carcinoma of upper inner quadrant of right female breast |
| 36712723 | Infiltrating ductal carcinoma of upper outer quadrant of right female breast |
| 36712724 | Infiltrating lobular carcinoma of left female breast |
| 36712725 | Infiltrating lobular carcinoma of right female breast |
| 36717260 | Infiltrating ductal carcinoma of central portion of left female breast |
| 37016439 | Primary invasive malignant neoplasm of female breast |
| 37018660 | Primary malignant inflammatory neoplasm of female breast |
| 37208322 | Infiltrating ductal carcinoma of axillary tail of left female breast |
| 37208324 | Infiltrating ductal carcinoma of lower inner quadrant of left female breast |
| 37208325 | Infiltrating ductal carcinoma of lower outer quadrant of left female breast |
| 37208326 | Infiltrating ductal carcinoma of axillary tail of right female breast |
| 37208328 | Infiltrating ductal carcinoma of lower inner quadrant of right female breast |
| 37208329 | Infiltrating ductal carcinoma of lower outer quadrant of right female breast |
| 40486565 | Sarcoma of axillary tail of female breast |
| 40489485 | Sarcoma of lower outer quadrant of female breast |
| 40489897 | Sarcoma of upper inner quadrant of female breast |
| 40489927 | Sarcoma lower inner quadrant of female breast |
| 40489928 | Sarcoma of central portion of female breast |
| 40489942 | Sarcoma of female breast |
| 40489945 | Sarcoma upper outer quadrant of female breast |
| 40492507 | Infiltrating duct carcinoma of female breast |
| 42512247 | Plasmablastic lymphoma of nipple |
| 42512452 | HHV8 positive diffuse large B-cell lymphoma of nipple |
| 42512503 | Myofibroblastic sarcoma of nipple |
| 42512576 | ALK positive large B-cell lymphoma of nipple |
| 42512817 | Adenomyoepithelioma with carcinoma of nipple |
| 42512849 | Invasive micropapillary carcinoma of breast of nipple |
| 42512881 | Solid papillary carcinoma with invasion of nipple |
| 44499448 | Paget disease and infiltrating duct carcinoma of breast of nipple |
| 44499538 | Cribriform carcinoma, NOS, of nipple |
| 44499675 | Carcinoma, NOS, of nipple |

Table D3 – continued from previous page
| Concept ID | Concept name |
| --- | --- |
| 44499753 | Infiltrating duct mixed with other types of carcinoma of nipple |
| 44499790 | Papillary carcinoma, NOS, of nipple |
| 44499805 | Tubular adenocarcinoma of nipple |
| 44500370 | Paget disease and intraductal carcinoma of breast of nipple |
| 44500439 | Adenocarcinoma, NOS, of nipple |
| 44500441 | Scirrhous adenocarcinoma of nipple |
| 44500599 | Infiltrating duct carcinoma, NOS, of nipple |
| 44500882 | Lobular carcinoma, NOS, of nipple |
| 44500940 | Infiltrating duct and lobular carcinoma of nipple |
| 44501583 | Inflammatory carcinoma of nipple |
| 44501805 | Atypical medullary carcinoma of nipple |
| 44501894 | Mucinous adenocarcinoma of nipple |
| 133711 | Overlapping malignant neoplasm of female breast |
| 137809 | Primary malignant neoplasm of female breast |
| 432263 | Primary malignant neoplasm of lower inner quadrant of female breast |
| 432845 | Primary malignant neoplasm of central portion of female breast |
| 433148 | Primary malignant neoplasm of areola of female breast |
| 436353 | Primary malignant neoplasm of upper outer quadrant of female breast |
| 440956 | Primary malignant neoplasm of upper inner quadrant of female breast |
| 441515 | Primary malignant neoplasm of lower outer quadrant of female breast |
| 442127 | Primary malignant neoplasm of nipple of female breast |
| 765123 | Primary malignant neoplasm of female right breast |
| 4091464 | Malignant neoplasm of nipple and areola of female breast |
| 4091465 | Malignant neoplasm of upper-outer quadrant of female breast |
| 4091466 | Malignant neoplasm of lower-outer quadrant of female breast |
| 4091467 | Malignant neoplasm of axillary tail of female breast |
| 4092511 | Malignant neoplasm of central part of female breast |
| 4092512 | Malignant neoplasm of upper-inner quadrant of female breast |
| 4095740 | Malignant neoplasm of lower-inner quadrant of female breast |
| 4095741 | Malignant neoplasm of ectopic site of female breast |
| 4157332 | Malignant neoplasm of female breast |
| 4246036 | Primary malignant neoplasm of ectopic female breast tissue |
| 36684817 | Primary malignant neoplasm of axillary tail of left female breast |
| 36684818 | Primary malignant neoplasm of female left breast |
| 36684819 | Primary malignant neoplasm of central portion of female left breast |
| 36684820 | Primary malignant neoplasm of axillary tail of right female breast |
| 36684821 | Primary malignant neoplasm of central portion of female right breast |
| 36712738 | Recurrent primary malignant neoplasm of left female breast |
| 36712739 | Recurrent primary malignant neoplasm of right female breast |
| 36712934 | Primary malignant neoplasm of bilateral female breasts |
| 40486563 | Carcinoma of female breast |

**Table D4:** Metastatic or metastatic breast cancer concepts used to block controls.

| Concept ID | Concept name |
| --- | --- |
| 140960 | Metastatic malignant neoplasm of female breast |
| 4110019 | Metastasis to breast of unknown primary<sup>†</sup> (updated) |
| 4246350 | Metastatic malignant neoplasm of areola of female breast |
| 4246352 | Metastatic malignant neoplasm of axillary tail of female breast |
| 4248184 | Metastatic malignant neoplasm of nipple of female breast |
| 4312296 | Metastatic malignant neoplasm of ectopic female breast tissue |
| 4312544 | Metastatic malignant neoplasm of lower inner quadrant of female breast |
| 4312546 | Metastatic malignant neoplasm of lower outer quadrant of female breast |
| 4312938 | Metastatic malignant neoplasm of central portion of female breast |
| 4314072 | Metastatic malignant neoplasm of upper inner quadrant of female breast |
| 4314189 | Metastatic malignant neoplasm of upper outer quadrant of female breast |

#### D.3 Hypertension

**Table D5:** Essential-hypertension case concepts.

| Concept ID | Concept name |
| --- | --- |
| 312648 | Benign essential hypertension |
| 317898 | Malignant essential hypertension |
| 320128 | Essential hypertension |
| 4034031 | Benign essential hypertension complicating AND/OR reason for care during pregnancy |
| 4058987 | High-renin essential hypertension |
| 4083723 | Essential hypertension complicating AND/OR reason for care during childbirth |
| 4148205 | Benign essential hypertension complicating AND/OR reason for care during puerperium |
| 4159755 | Labile essential hypertension |
| 4180283 | Systolic essential hypertension |
| 4215640 | Benign essential hypertension complicating AND/OR reason for care during childbirth |
| 4217486 | Essential hypertension in obstetric context |
| 4263067 | Low-renin essential hypertension |
| 4269358 | Benign essential hypertension in obstetric context |
| 4302591 | Essential hypertension complicating AND/OR reason for care during pregnancy |
| 4321603 | Essential hypertension complicating AND/OR reason for care during puerperium |
| 45757787 | Postpartum pre-existing essential hypertension |

**Table D6:** Hypertension control-blocking concepts.

| Concept ID | Concept name |
| --- | --- |
| 134414 | Pre-eclampsia or eclampsia with pre-existing hypertension – delivered with postnatal complication<sup>†</sup> (deprecated) |
| 135601 | Pre-eclampsia or eclampsia with pre-existing hypertension – delivered<sup>†</sup> (deprecated) |
| 136743 | Pre-eclampsia or eclampsia with pre-existing hypertension – not delivered<sup>†</sup> (deprecated) |
| 141084 | Pre-eclampsia or eclampsia with pre-existing hypertension |
| 192684 | Renal hypertension complicating pregnancy, childbirth and the puerperium – not delivered<sup>†</sup> (deprecated) |
| 197930 | Renal hypertension complicating pregnancy, childbirth and the puerperium |
| 200157 | Renal hypertension complicating pregnancy, childbirth and the puerperium – delivered<sup>†</sup> (deprecated) |
| 201912 | Renal hypertension complicating pregnancy, childbirth and the puerperium – delivered with postnatal complication<sup>†</sup> (deprecated) |

Table D6 – continued from previous page
| Concept ID | Concept name |
| --- | --- |
| 312648 | Benign essential hypertension |
| 314103 | Benign essential hypertension complicating pregnancy, childbirth and the puerperium – delivered<sup>†</sup> (deprecated) |
| 314423 | Benign essential hypertension complicating pregnancy, childbirth and the puerperium – not delivered<sup>†</sup> (deprecated) |
| 314958 | Benign secondary hypertension |
| 316866 | Hypertensive disorder |
| 317895 | Renovascular hypertension |
| 317898 | Malignant essential hypertension |
| 318437 | Malignant secondary hypertension |
| 319826 | Secondary hypertension |
| 320128 | Essential hypertension |
| 320456 | Benign essential hypertension complicating pregnancy, childbirth and the puerperium – delivered with postnatal complication<sup>†</sup> (deprecated) |
| 321074 | Pre-existing hypertension complicating pregnancy, childbirth and puerperium |
| 321080 | Hypertension complicating pregnancy, childbirth and the puerperium |
| 321638 | Benign essential hypertension complicating pregnancy, childbirth and the puerperium |
| 443771 | Renal hypertension |
| 1340364 | Exacerbation of hypertensive disorder |
| 3169253 | Hypertension with apparent mineralocorticoid excess |
| 3191244 | Hypertension with hyperadrenergic findings |
| 3656115 | Hypertension due to aortic arch obstruction |
| 4006325 | Hypertension secondary to renal disease in obstetric context |
| 4023318 | Malignant hypertension complicating AND/OR reason for care during childbirth |
| 4028741 | Benign hypertension |
| 4028951 | Malignant hypertension complicating AND/OR reason for care during puerperium |
| 4032952 | Renal sclerosis with hypertension |
| 4034031 | Benign essential hypertension complicating AND/OR reason for care during pregnancy |
| 4048212 | Goldblatt hypertension |
| 4049389 | Paroxysmal hypertension |
| 4057978 | Pre-existing hypertensive heart and renal disease complicating pregnancy, childbirth and the puerperium |
| 4057979 | Pre-existing secondary hypertension complicating pregnancy, childbirth and puerperium |
| 4058987 | High-renin essential hypertension |
| 4061667 | Hypertension induced by oral contraceptive pill |
| 4062550 | Pre-eclampsia or eclampsia with pre-existing hypertension with postnatal complication |
| 4062811 | Benign essential hypertension complicating pregnancy, childbirth and the puerperium with postnatal complication |

Table D6 – continued from previous page
| Concept ID | Concept name |
| --- | --- |
| 4080325 | Chronic hypertension complicating AND/OR reason for care during puerperium |
| 4083723 | Essential hypertension complicating AND/OR reason for care during childbirth |
| 4094374 | Hypertension secondary to renal disease complicating AND/OR reason for care during childbirth |
| 4108213 | Hypertension secondary to drug |
| 4110947 | Malignant secondary renovascular hypertension |
| 4110948 | Hypertension secondary to endocrine disorder |
| 4118910 | Maternal hypertension |
| 4146816 | Pre-existing hypertension complicating AND/OR reason for care during childbirth |
| 4148205 | Benign essential hypertension complicating AND/OR reason for care during puerperium |
| 4151903 | Pre-existing hypertension complicating AND/OR reason for care during puerperium |
| 4159755 | Labile essential hypertension |
| 4162306 | Malignant hypertension complicating AND/OR reason for care during pregnancy |
| 4167358 | Diastolic hypertension |
| 4174979 | Hypertension secondary to renal disease complicating AND/OR reason for care during pregnancy |
| 4178312 | Hypertension secondary to kidney transplant |
| 4180283 | Systolic essential hypertension |
| 4209293 | Systolic hypertension |
| 4212496 | Parenchymal renal hypertension |
| 4215640 | Benign essential hypertension complicating AND/OR reason for care during childbirth |
| 4217486 | Essential hypertension in obstetric context |
| 4218088 | Malignant hypertension in obstetric context |
| 4219323 | Hypertension secondary to renal disease complicating AND/OR reason for care during puerperium |
| 4221991 | Rebound hypertension |
| 4227607 | Chronic hypertension in obstetric context |
| 4242878 | Sustained diastolic hypertension |
| 4249016 | Benign secondary renovascular hypertension |
| 4253928 | Secondary diastolic hypertension |
| 4262182 | Hypertension with albuminuria |
| 4263067 | Low-renin essential hypertension |
| 4269358 | Benign essential hypertension in obstetric context |
| 4276511 | Labile diastolic hypertension |
| 4277110 | Pre-existing hypertension complicating AND/OR reason for care during pregnancy |
| 4279525 | Hypertension in the obstetric context |
| 4283352 | Pre-eclampsia added to pre-existing hypertension |

Table D6 – continued from previous page
| Concept ID | Concept name |
| --- | --- |
| 4289142 | Eclampsia added to pre-existing hypertension |
| 4289933 | Malignant hypertension |
| 4291933 | Chronic hypertension complicating AND/OR reason for care during pregnancy |
| 4302591 | Essential hypertension complicating AND/OR reason for care during pregnancy |
| 4304837 | Chronic hypertension complicating AND/OR reason for care during childbirth |
| 4305599 | Renal arterial hypertension |
| 4311246 | Pre-existing hypertension in obstetric context |
| 4321603 | Essential hypertension complicating AND/OR reason for care during puerperium |
| 4322735 | Hypertension associated with transplantation |
| 35624277 | Hypertension due to gain-of-function mutation in mineralocorticoid receptor |
| 36713024 | Labile systemic arterial hypertension |
| 36715087 | Brachydactyly and arterial hypertension syndrome |
| 37016726 | Supine hypertension |
| 37208172 | Resistant hypertensive disorder |
| 40481896 | Hypertensive urgency |
| 42538697 | Diastolic hypertension and systolic hypertension |
| 42538946 | Hypertension complicating pregnancy |
| 42873163 | Hypertension complicating pregnancy, childbirth and the puerperium, antepartum |
| 43020424 | Hypertensive emergency |
| 44783643 | Intermittent hypertension |
| 44784484 | Hypertension in the puerperium with pulmonary edema |
| 44809026 | Stage 1 hypertension (NICE – National Institute for Health and Clinical Excellence 2011) |
| 44809027 | Severe hypertension (NICE – National Institute for Health and Clinical Excellence 2011) |
| 44809548 | Hypertension resistant to drug therapy |
| 44809569 | Stage 2 hypertension (NICE – National Institute for Health and Clinical Excellence 2011) |
| 44811110 | Nocturnal hypertension |
| 44811932 | Stage 1 hypertension (NICE 2011) without evidence of end organ damage |
| 44811933 | Stage 1 hypertension (NICE 2011) with evidence of end organ damage |
| 45757119 | Eclampsia with pre-existing hypertension in childbirth |
| 45757137 | Pre-existing hypertensive heart and chronic kidney disease in mother complicating childbirth |
| 45757356 | Pre-existing hypertensive chronic kidney disease in mother complicating pregnancy |
| 45757444 | Hypertension in chronic kidney disease stage 5 due to type 2 diabetes mellitus |
| 45757445 | Hypertension in chronic kidney disease stage 4 due to type 2 diabetes mellitus |
| 45757446 | Hypertension in chronic kidney disease stage 3 due to type 2 diabetes mellitus |
| 45757447 | Hypertension in chronic kidney disease stage 2 due to type 2 diabetes mellitus |
| 45757756 | Page kidney |

Table D6 – continued from previous page
| Concept ID | Concept name |
| --- | --- |
| 45757787 | Postpartum pre-existing essential hypertension |
| 45768449 | Hypertensive crisis |
| 45771064 | Hypertension in chronic kidney disease due to type 2 diabetes mellitus |
| 45771067 | Hypertension in chronic kidney disease due to type 1 diabetes mellitus |

#### D.4 Inflammatory bowel disease

**Table D7:** Inflammatory bowel disease case concepts.

| Concept ID | Concept name |
| --- | --- |
| 77317 | Chronic ulcerative rectosigmoiditis |
| 78799 | Chronic ulcerative ileocolitis |
| 81893 | Ulcerative colitis |
| 194077 | Chronic ulcerative enterocolitis |
| 194684 | Crohn's disease of large bowel |
| 195575 | Crohn's disease of small and large intestines |
| 195585 | Crohn's disease of small intestine |
| 201606 | Crohn's disease |
| 602593 | Acute ulcerative pancolitis |
| 602594 | Acute left-sided ulcerative colitis |
| 602607 | Acute ulcerative rectosigmoiditis |
| 1340297 | Exacerbation of Crohn's disease |
| 1340490 | Exacerbation of ulcerative colitis |
| 1340491 | Exacerbation of ulcerative proctocolitis |
| 3184226 | Pancolonic ulcerative colitis |
| 4025853 | Ulcerative ileocolitis |
| 4029372 | Acute ulcerative colitis |
| 4031048 | Severe chronic ulcerative colitis |
| 4055020 | Crohn's disease of terminal ileum |
| 4055884 | Crohn's disease of oral soft tissues |
| 4086978 | Eosinophilic ulcerative colitis |
| 4122617 | Crohn's disease of gingivae |
| 4142544 | Crohn's disease in remission |
| 4166480 | Ulcerative proctocolitis |
| 4177488 | Crohn's disease of colon |
| 4187900 | Exacerbation of ulcerative colitis |
| 4210469 | Crohn's disease of duodenum |
| 4212991 | Exacerbation of Crohn's disease of large intestine |
| 4212992 | Exacerbation of Crohn's disease of small intestine |
| 4239382 | Crohn's disease of jejunum |
| 4242392 | Crohn's disease of rectum |
| 4244235 | Crohn's disease of ileum |
| 4246693 | Crohn's disease of intestine |
| 4259504 | Iritis with ulcerative colitis |
| 4264850 | Gastrointestinal Crohn's disease |
| 4266370 | Crohn's disease of pylorus |
| 4301738 | Mild chronic ulcerative colitis |
| 4302002 | Moderate chronic ulcerative colitis |
| 4323289 | Crohn's disease of pyloric antrum |
| 4340114 | Crohn's disease of esophagus |
| 4341633 | Indeterminate colitis |

Table D7 – continued from previous page
| Concept ID | Concept name |
| --- | --- |
| 4342643 | Crohn's disease of stomach |
| 4342656 | Ulcerative enterocolitis |
| 36686095 | Crohn's disease of small intestine with stenosis |
| 36715915 | Crohn disease of anal canal |
| 36716695 | Crohn's disease of gastrointestinal anastomosis |
| 36716986 | Arthritis co-occurrent and due to Crohn's disease |
| 37116446 | Crohn disease of appendix |
| 40479837 | Chronic ulcerative colitis |
| 40479839 | Ulcerative pancolitis |
| 40481367 | Chronic left-sided ulcerative colitis |
| 40482241 | Chronic ulcerative pancolitis |
| 40482865 | Left sided ulcerative colitis |
| 42537666 | Crohn disease of upper gastrointestinal tract |
| 44783784 | Ulcerative colitis in remission |
| 46269838 | Abscess of intestine co-occurrent and due to chronic ulcerative pancolitis |
| 46269848 | Abscess of intestine co-occurrent and due to chronic ulcerative rectosigmoiditis |
| 46269878 | Abscess of intestine co-occurrent and due to Crohn's disease of small and large intestine |
| 46269883 | Abscess of intestine co-occurrent and due to Crohn's disease of small intestine |
| 46269888 | Abscess of intestine co-occurrent and due to Crohn's disease |
| 46269951 | Abscess of intestine co-occurrent and due to ulcerative colitis |
| 46274073 | Abscess of intestine co-occurrent and due to Crohn's disease of large intestine |

**Table D8:** Inflammatory bowel disease control-exclusion concepts.

| Concept ID | Concept name |
| --- | --- |
| 192960 | Noninfectious gastroenteritis |
| 201773 | Enteritis of small intestine |
| 443865 | Noninfectious colitis |
| 4074815 | Inflammatory bowel disease |
| 4083999 | Noninfectious jejunitis |
| 4145072 | Noninfective enteritis and colitis |
| 4196810 | Noninfectious ileitis |
| 4201583 | Noninfectious sigmoiditis |
| 4234755 | Idiopathic chronic inflammatory bowel disease<sup>†</sup> (updated) |
| 4272488 | Colitis |
| 4324838 | Noninfectious enteritis of intestine |

#### D.5 Melanoma

**Table D9:** Invasive cutaneous melanoma case concepts.

| Concept ID | Concept name |
| --- | --- |
| 133419 | Overlapping malignant melanoma of skin |
| 133713 | Malignant melanoma of skin of face |
| 133714 | Malignant melanoma of skin of trunk |
| 134294 | Malignant melanoma of skin of external auditory canal |
| 139757 | Malignant melanoma of skin of lower limb |
| 140048 | Malignant melanoma of skin of lip |
| 141232 | Malignant melanoma of skin |
| 434590 | Malignant melanoma of skin of eyelid |
| 438983 | Malignant melanoma of skin of upper limb |
| 440050 | Malignant melanoma of skin of neck |
| 442143 | Malignant melanoma of skin of scalp |
| 444462 | Malignant melanoma of skin of ear |
| 602586 | Malignant melanoma of skin of right ankle |
| 602588 | Malignant melanoma of skin of right wrist |
| 602597 | Malignant melanoma of skin of left wrist |
| 604286 | Malignant melanoma of skin of left forearm |
| 608893 | Bilateral malignant melanoma of skin of lower limbs |
| 608894 | Bilateral malignant melanoma of skin of upper limbs |
| 616697 | Malignant melanoma of skin of left ankle |
| 619931 | Malignant melanoma of skin of right forearm |
| 3183080 | Malignant melanoma of skin with BRAF V600E sequence variant |
| 4080901 | Amelanotic malignant melanoma of skin |
| 4092364 | Malignant melanoma of auricle (ear) |
| 4095591 | Malignant melanoma of temple<sup>†</sup> (updated) |
| 4110725 | Superficial spreading malignant melanoma of skin |
| 4110726 | Acral lentiginous malignant melanoma of skin |
| 4110727 | Malignant melanoma arising in intradermal nevus |
| 4112760 | Nodular malignant melanoma of skin |
| 4115298 | Malignant melanoma arising in congenital nevus |
| 4116198 | Lentigo maligna melanoma |
| 4141249 | Malignant blue nevus of skin |
| 4244049 | Malignant melanoma of skin of abdomen |
| 4244050 | Malignant melanoma of skin of back |
| 4244051 | Malignant melanoma of skin of breast |
| 4244166 | Malignant melanoma of skin of cheek |
| 4244167 | Malignant melanoma of skin of chin |
| 4244168 | Malignant melanoma of skin of finger |
| 4244169 | Malignant melanoma of skin of foot |
| 4244170 | Malignant melanoma of skin of forearm |
| 4244488 | Malignant melanoma of skin of nose |
| 4244489 | Malignant melanoma of skin of perineum |

Table D9 – continued from previous page
| Concept ID | Concept name |
| --- | --- |
| 4245465 | Malignant melanoma of skin of ankle |
| 4245466 | Malignant melanoma of skin of axilla |
| 4245467 | Malignant melanoma of skin of buttock |
| 4245918 | Malignant melanoma of skin of eyebrow |
| 4245919 | Malignant melanoma of skin of groin |
| 4246470 | Malignant melanoma of perianal skin |
| 4246472 | Malignant melanoma of skin of chest |
| 4246473 | Malignant melanoma of skin of elbow |
| 4246474 | Malignant melanoma of skin of forehead |
| 4246790 | Malignant melanoma of skin of hip |
| 4246791 | Malignant melanoma of skin of popliteal area |
| 4246792 | Malignant melanoma of skin of temporal region |
| 4247221 | Malignant melanoma of skin of hand |
| 4247224 | Malignant melanoma of skin of thigh |
| 4247225 | Malignant melanoma of skin of umbilicus |
| 4247226 | Malignant melanoma of skin of wrist |
| 4247713 | Malignant melanoma of skin of knee |
| 4247714 | Malignant melanoma of skin of shoulder |
| 4247715 | Malignant melanoma of skin of toe |
| 4300096 | PUVA therapy-associated malignant melanoma |
| 4309088 | Malignant melanoma of skin of upper lip |
| 4309539 | Malignant melanoma of skin of lower eyelid |
| 4310839 | Malignant melanoma of skin of lower lip |
| 4313171 | Malignant melanoma of skin of canthus of eye |
| 4313205 | Malignant melanoma of skin of upper eyelid |
| 35617772 | Malignant melanoma of skin of left upper limb |
| 36518338 | Malignant melanoma in precancerous melanosis of skin of trunk |
| 36519380 | Malignant melanoma arising in giant congenital nevus of skin of lower limb and hip |
| 36519480 | Malignant melanoma in junctional nevus of skin of lower limb and hip |
| 36521362 | Acral melanoma of skin of lip, NOS |
| 36521679 | Malignant melanoma arising in giant congenital nevus of skin, NOS |
| 36521926 | Amelanotic melanoma of skin of lower limb and hip |
| 36522683 | Balloon cell melanoma of skin of lower limb and hip |
| 36523026 | Malignant melanoma, regressing of skin of lower limb and hip |
| 36524101 | Malignant melanoma, regressing of skin of other and unspecified parts of face |
| 36525177 | Nodular melanoma of skin of upper limb and shoulder |
| 36525199 | Malignant melanoma, regressing of external ear |
| 36526412 | Acral melanoma of eyelid |
| 36526559 | Amelanotic melanoma of skin of upper limb and shoulder |
| 36526622 | Acral melanoma of overlapping lesion of skin |
| 36527048 | Balloon cell melanoma of skin, NOS |
| 36527205 | Malignant melanoma, regressing of skin of upper limb and shoulder |
| 36528293 | Balloon cell melanoma of external ear |

Table D9 – continued from previous page
| Concept ID | Concept name |
| --- | --- |
| 36529065 | Malignant melanoma, regressing of eyelid |
| 36529357 | Blue nevus, malignant of skin of other and unspecified parts of face |
| 36529405 | Lentigo maligna melanoma of overlapping lesion of skin |
| 36532308 | Malignant melanoma arising in giant congenital nevus of skin of upper limb and shoulder |
| 36532847 | Malignant melanoma in junctional nevus of eyelid |
| 36533016 | Malignant melanoma in junctional nevus of skin, NOS |
| 36533239 | Blue nevus, malignant of overlapping lesion of skin |
| 36533686 | Blue nevus, malignant of skin of upper limb and shoulder |
| 36534975 | Balloon cell melanoma of overlapping lesion of skin |
| 36535699 | Lentigo maligna melanoma of skin of lip, NOS |
| 36535705 | Amelanotic melanoma of skin of lip, NOS |
| 36536386 | Blue nevus, malignant of skin of scalp and neck |
| 36536415 | Lentigo maligna melanoma of skin of lower limb and hip |
| 36536797 | Malignant Spitz tumor of skin of trunk |
| 36537261 | Desmoplastic melanoma, NOS, of skin of lower limb and hip |
| 36538436 | Desmoplastic melanoma, NOS, of overlapping lesion of skin |
| 36538652 | Malignant melanoma in junctional nevus of overlapping lesion of skin |
| 36538697 | Amelanotic melanoma of external ear |
| 36538883 | Malignant melanoma arising in giant congenital nevus of skin of lip, NOS |
| 36538971 | Balloon cell melanoma of skin of upper limb and shoulder |
| 36539718 | Malignant melanoma in precancerous melanosis of skin of lower limb and hip |
| 36540636 | Desmoplastic melanoma, NOS, of skin of lip, NOS |
| 36540643 | Malignant melanoma in precancerous melanosis of overlapping lesion of skin |
| 36542886 | Epithelioid cell melanoma of overlapping lesion of skin |
| 36543197 | Lentigo maligna melanoma of skin of upper limb and shoulder |
| 36543265 | Malignant melanoma in precancerous melanosis of external ear |
| 36543603 | Malignant melanoma arising in giant congenital nevus of skin of scalp and neck |
| 36544050 | Balloon cell melanoma of eyelid |
| 36544667 | Desmoplastic melanoma, NOS, of skin of upper limb and shoulder |
| 36544787 | Spindle cell melanoma, NOS, of skin of lower limb and hip |
| 36545191 | Nodular melanoma of skin of lower limb and hip |
| 36545723 | Malignant melanoma, NOS, of overlapping lesion of skin |
| 36545835 | Epithelioid cell melanoma of skin of lip, NOS |
| 36545924 | Low cumulative sun damage melanoma of skin of lip, NOS |
| 36546325 | Balloon cell melanoma of skin of other and unspecified parts of face |
| 36546648 | Malignant Spitz tumor of skin of lip, NOS |
| 36547183 | Low cumulative sun damage melanoma of overlapping lesion of skin |
| 36547254 | Blue nevus, malignant of skin of lower limb and hip |
| 36549146 | Amelanotic melanoma of overlapping lesion of skin |
| 36549966 | Malignant Spitz tumor of skin of lower limb and hip |
| 36550489 | Malignant melanoma in precancerous melanosis of skin of other and unspecified parts of face |

Table D9 – continued from previous page
| Concept ID | Concept name |
| --- | --- |
| 36551092 | Balloon cell melanoma of skin of lip, NOS |
| 36551392 | Low cumulative sun damage melanoma of skin of lower limb and hip |
| 36551591 | Malignant melanoma in junctional nevus of skin of lip, NOS |
| 36552402 | Malignant melanoma, NOS, of skin of upper limb and shoulder |
| 36552448 | Blue nevus, malignant of skin of lip, NOS |
| 36553353 | Low cumulative sun damage melanoma of skin of upper limb and shoulder |
| 36555212 | Malignant melanoma in precancerous melanosis of skin of scalp and neck |
| 36555693 | Malignant melanoma in junctional nevus of skin of other and unspecified parts of face |
| 36555705 | Malignant Spitz tumor of skin of upper limb and shoulder |
| 36556372 | Epithelioid cell melanoma of skin of other and unspecified parts of face |
| 36556552 | Balloon cell melanoma of skin of scalp and neck |
| 36557043 | Malignant melanoma arising in giant congenital nevus of overlapping lesion of skin |
| 36557414 | Acral melanoma of external ear |
| 36558120 | Acral melanoma of skin of upper limb and shoulder |
| 36559367 | Malignant melanoma, regressing of skin of lip, NOS |
| 36559388 | Amelanotic melanoma of eyelid |
| 36559573 | Malignant melanoma, regressing of overlapping lesion of skin |
| 36559731 | Epithelioid cell melanoma of skin, NOS |
| 36559746 | Epithelioid cell melanoma of eyelid |
| 36559941 | Spindle cell melanoma, NOS, of skin of upper limb and shoulder |
| 36561208 | Epithelioid cell melanoma of skin of upper limb and shoulder |
| 36561552 | Spindle cell melanoma, NOS, of overlapping lesion of skin |
| 36563329 | Malignant melanoma in junctional nevus of skin of scalp and neck |
| 36563439 | Acral melanoma of skin of lower limb and hip |
| 36564259 | Malignant melanoma in precancerous melanosis of skin of upper limb and shoulder |
| 36564461 | Spindle cell melanoma, NOS, of eyelid |
| 36564549 | Malignant melanoma, regressing of skin, NOS |
| 36564904 | Malignant melanoma arising in giant congenital nevus of external ear |
| 36565129 | Malignant melanoma in junctional nevus of skin of upper limb and shoulder |
| 36565165 | Acral melanoma of skin of scalp and neck |
| 36565315 | Malignant melanoma in precancerous melanosis of skin of lip, NOS |
| 36565542 | Malignant Spitz tumor of skin of scalp and neck |
| 36565544 | Malignant melanoma in precancerous melanosis of eyelid |
| 36566011 | Epithelioid cell melanoma of skin of lower limb and hip |
| 36566338 | Malignant melanoma in precancerous melanosis of skin, NOS |
| 36566436 | Malignant Spitz tumor of overlapping lesion of skin |
| 36566825 | Malignant melanoma arising in giant congenital nevus of eyelid |
| 36567403 | Desmoplastic melanoma, NOS, of eyelid |
| 36567448 | Nodular melanoma of overlapping lesion of skin |
| 36712733 | Malignant melanoma of skin of left lower limb |
| 37111319 | Familial malignant melanoma of skin |

Table D9 – continued from previous page
| Concept ID | Concept name |
| --- | --- |
| 37208039 | Malignant melanoma of skin of right upper limb |
| 40486090 | Malignant melanoma of skin of anus |
| 40488963 | Malignant melanoma of skin of scrotum |
| 40488990 | Malignant melanoma of skin of penis |
| 40488993 | Malignant melanoma of skin of vulva |
| 42535535 | Malignant melanoma of skin of right lower limb |
| 42709764 | Malignant melanoma of skin of lower leg |
| 42709765 | Malignant melanoma of skin of upper arm |
| 44499518 | Nodular melanoma of skin of trunk |
| 44499825 | Desmoplastic melanoma, NOS, of skin of other and unspecified parts of face |
| 44499826 | Epithelioid cell melanoma of skin of scalp and neck |
| 44499924 | Amelanotic melanoma of skin of other and unspecified parts of face |
| 44499925 | Malignant melanoma in junctional nevus of external ear |
| 44500152 | Malignant melanoma arising in giant congenital nevus of skin of trunk |
| 44500209 | Lentigo maligna melanoma of skin of scalp and neck |
| 44500263 | Spindle cell melanoma, NOS, of skin of trunk |
| 44500309 | Balloon cell melanoma of skin of trunk |
| 44500310 | Amelanotic melanoma of skin of trunk |
| 44500373 | Low cumulative sun damage melanoma of external ear |
| 44500512 | Spindle cell melanoma, NOS, of skin of lip, NOS |
| 44500607 | Desmoplastic melanoma, NOS, of skin of scalp and neck |
| 44500608 | Spindle cell melanoma, NOS, of skin of scalp and neck |
| 44500678 | Desmoplastic melanoma, NOS, of external ear |
| 44500740 | Malignant melanoma, regressing of skin of scalp and neck |
| 44500741 | Lentigo maligna melanoma of skin of other and unspecified parts of face |
| 44500742 | Desmoplastic melanoma, NOS, of skin of trunk |
| 44500804 | Malignant Spitz tumor of skin, NOS |
| 44501098 | Spindle cell melanoma, NOS, of skin, NOS |
| 44501099 | Blue nevus, malignant of skin of trunk |
| 44501359 | Low cumulative sun damage melanoma of skin of other and unspecified parts of face |
| 44501418 | Spindle cell melanoma, NOS, of skin of other and unspecified parts of face |
| 44501532 | Malignant melanoma, NOS, of external ear |
| 44501662 | Nodular melanoma of skin of lip, NOS |
| 44501663 | Nodular melanoma of external ear |
| 44501808 | Lentigo maligna melanoma of external ear |
| 44501809 | Acral melanoma of skin of other and unspecified parts of face |
| 44501856 | Low cumulative sun damage melanoma of skin of scalp and neck |
| 44501961 | Malignant melanoma, regressing of skin of trunk |
| 44501965 | Epithelioid cell melanoma of external ear |
| 44502043 | Acral melanoma of skin of trunk |
| 44502119 | Malignant melanoma, NOS, of skin of scalp and neck |
| 44502120 | Nodular melanoma of skin of scalp and neck |
| 44502124 | Malignant Spitz tumor of skin of other and unspecified parts of face |

Table D9 – continued from previous page
| Concept ID | Concept name |
| --- | --- |
| 44502294 | Lentigo maligna melanoma of eyelid |
| 44502400 | Amelanotic melanoma of skin of scalp and neck |
| 44502719 | Nodular melanoma of eyelid |
| 44502888 | Nodular melanoma of skin of other and unspecified parts of face |
| 44502889 | Low cumulative sun damage melanoma of eyelid |
| 44502964 | Malignant melanoma in junctional nevus of skin of trunk |
| 44502965 | Low cumulative sun damage melanoma of skin of trunk |
| 44503100 | Lentigo maligna melanoma of skin of trunk |
| 44503101 | Malignant melanoma arising in giant congenital nevus of skin of other and unspecified parts of face |
| 44503102 | Epithelioid cell melanoma of skin of trunk |
| 44503103 | Spindle cell melanoma, NOS, of external ear |
| 44505119 | Blue nevus, malignant of skin, NOS |
| 44505544 | Malignant melanoma, NOS, of skin of other and unspecified parts of face |

#### D.6 Multiple sclerosis

**Table D10:**
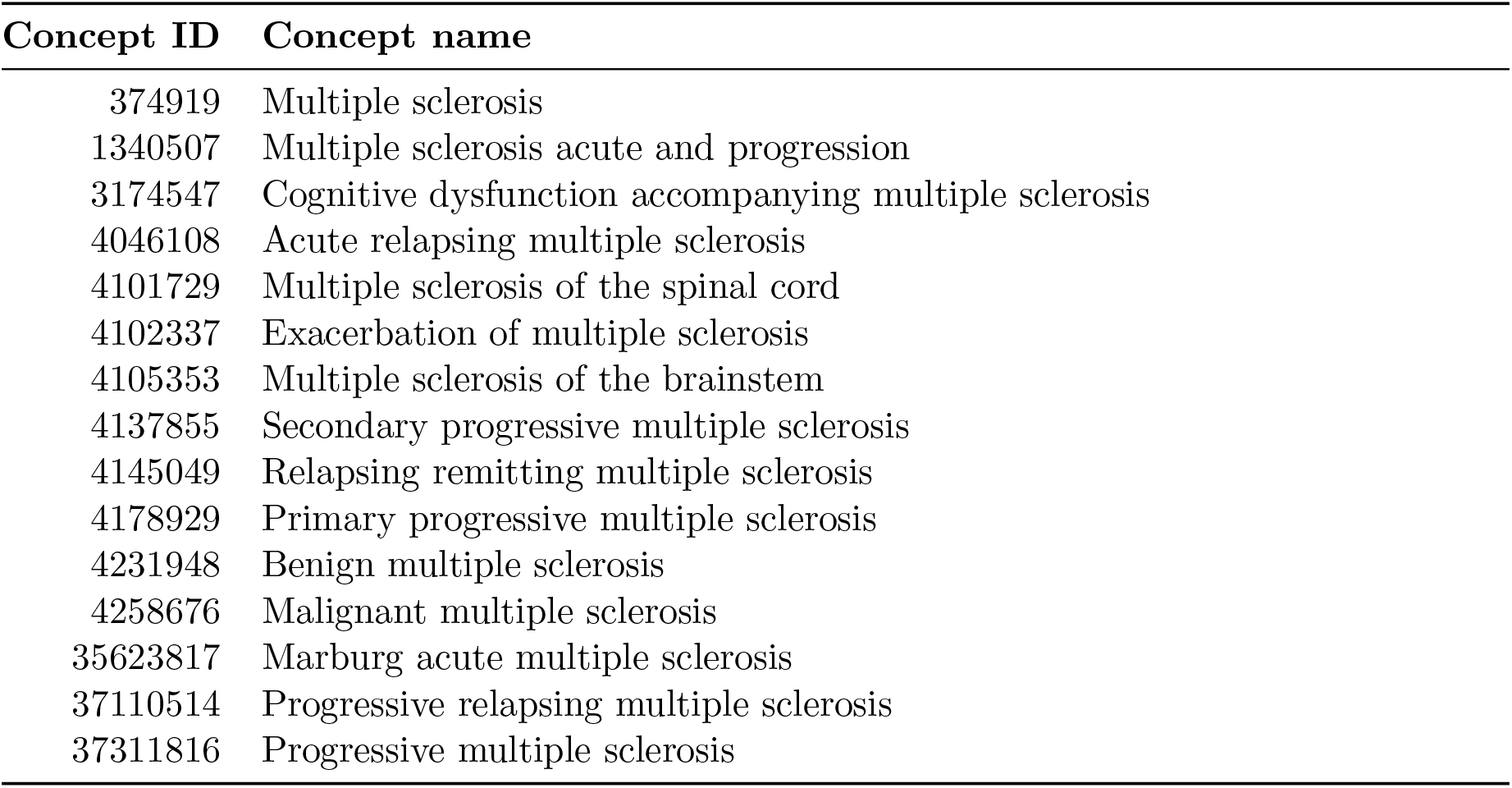
Multiple sclerosis case concepts.

| Concept ID | Concept name |
| --- | --- |
| 374919 | Multiple sclerosis |
| 1340507 | Multiple sclerosis acute and progression |
| 3174547 | Cognitive dysfunction accompanying multiple sclerosis |
| 4046108 | Acute relapsing multiple sclerosis |
| 4101729 | Multiple sclerosis of the spinal cord |
| 4102337 | Exacerbation of multiple sclerosis |
| 4105353 | Multiple sclerosis of the brainstem |
| 4137855 | Secondary progressive multiple sclerosis |
| 4145049 | Relapsing remitting multiple sclerosis |
| 4178929 | Primary progressive multiple sclerosis |
| 4231948 | Benign multiple sclerosis |
| 4258676 | Malignant multiple sclerosis |
| 35623817 | Marburg acute multiple sclerosis |
| 37110514 | Progressive relapsing multiple sclerosis |
| 37311816 | Progressive multiple sclerosis |

#### D.7 Prostate cancer

**Table D11:**
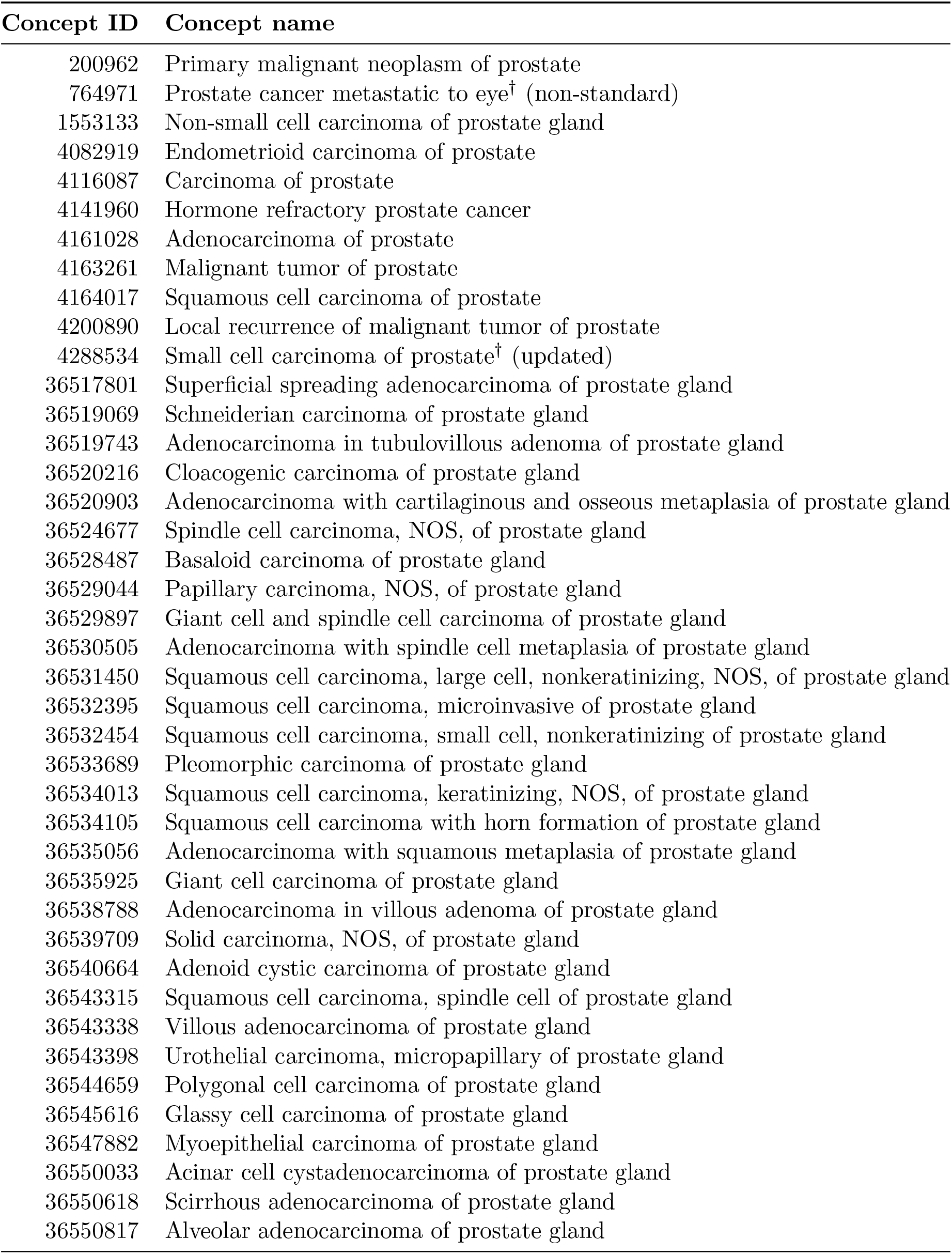
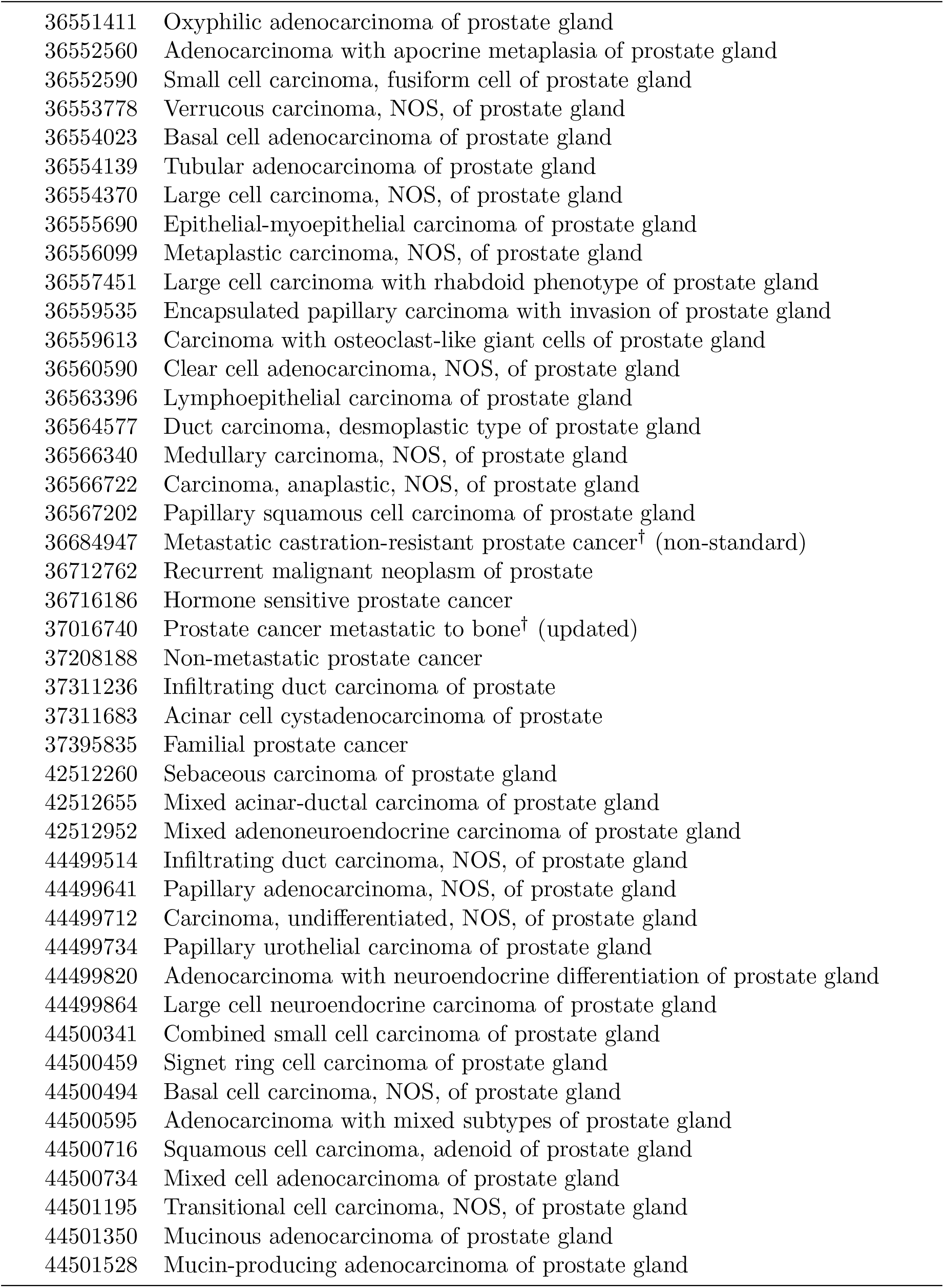
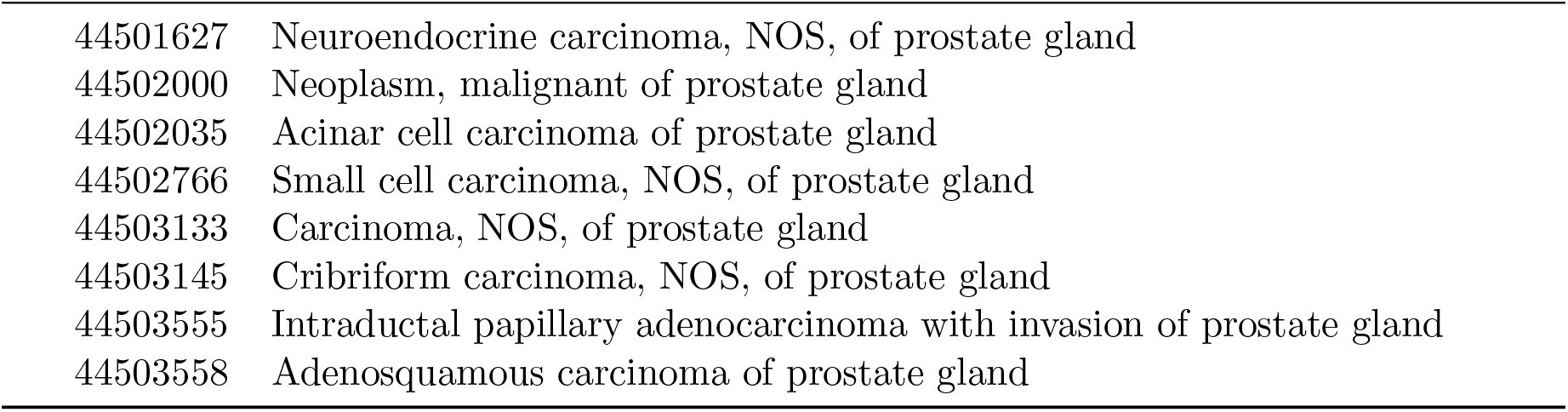
Prostate cancer direct and advanced case concepts.

**Table D12:**
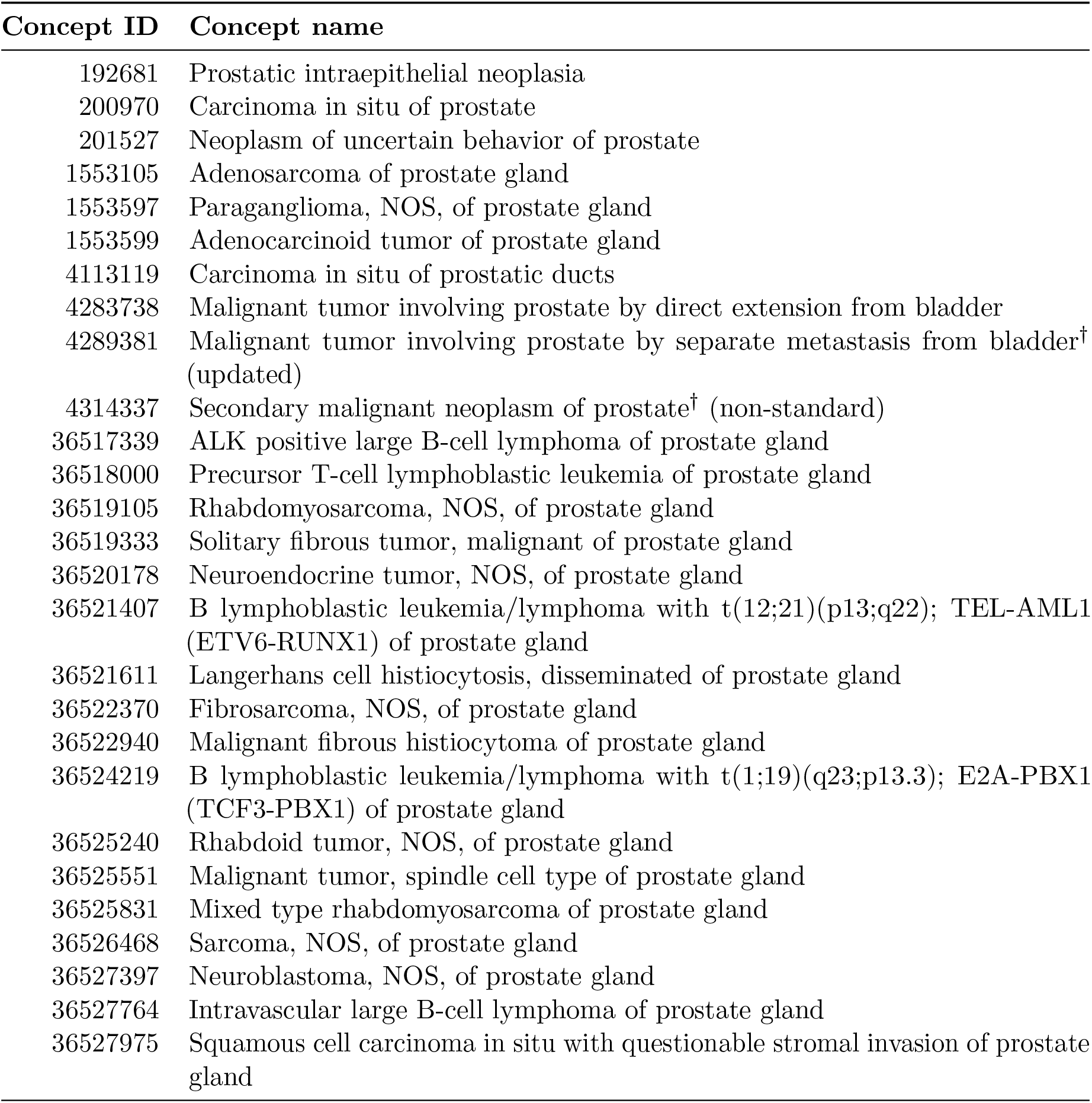
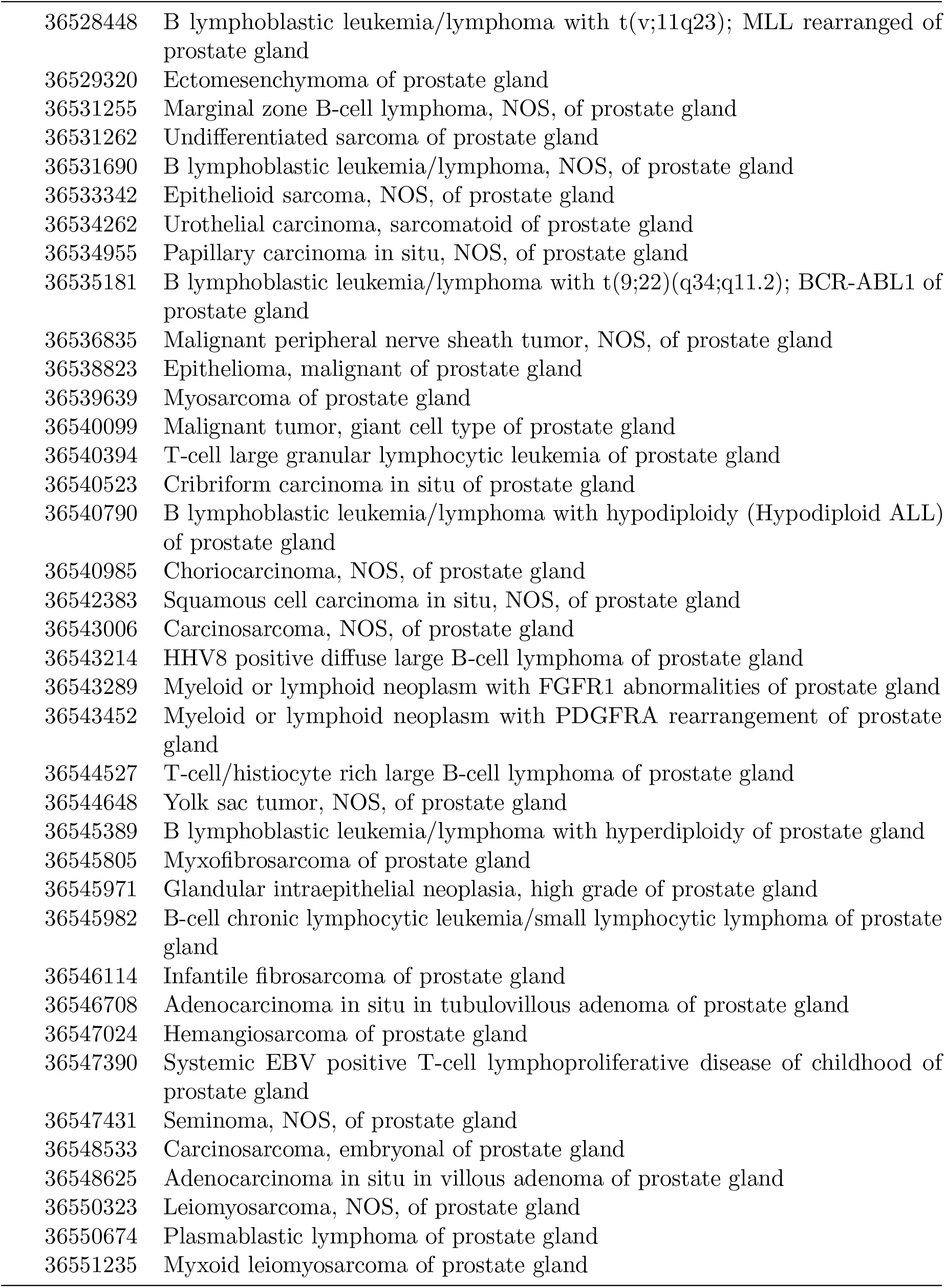
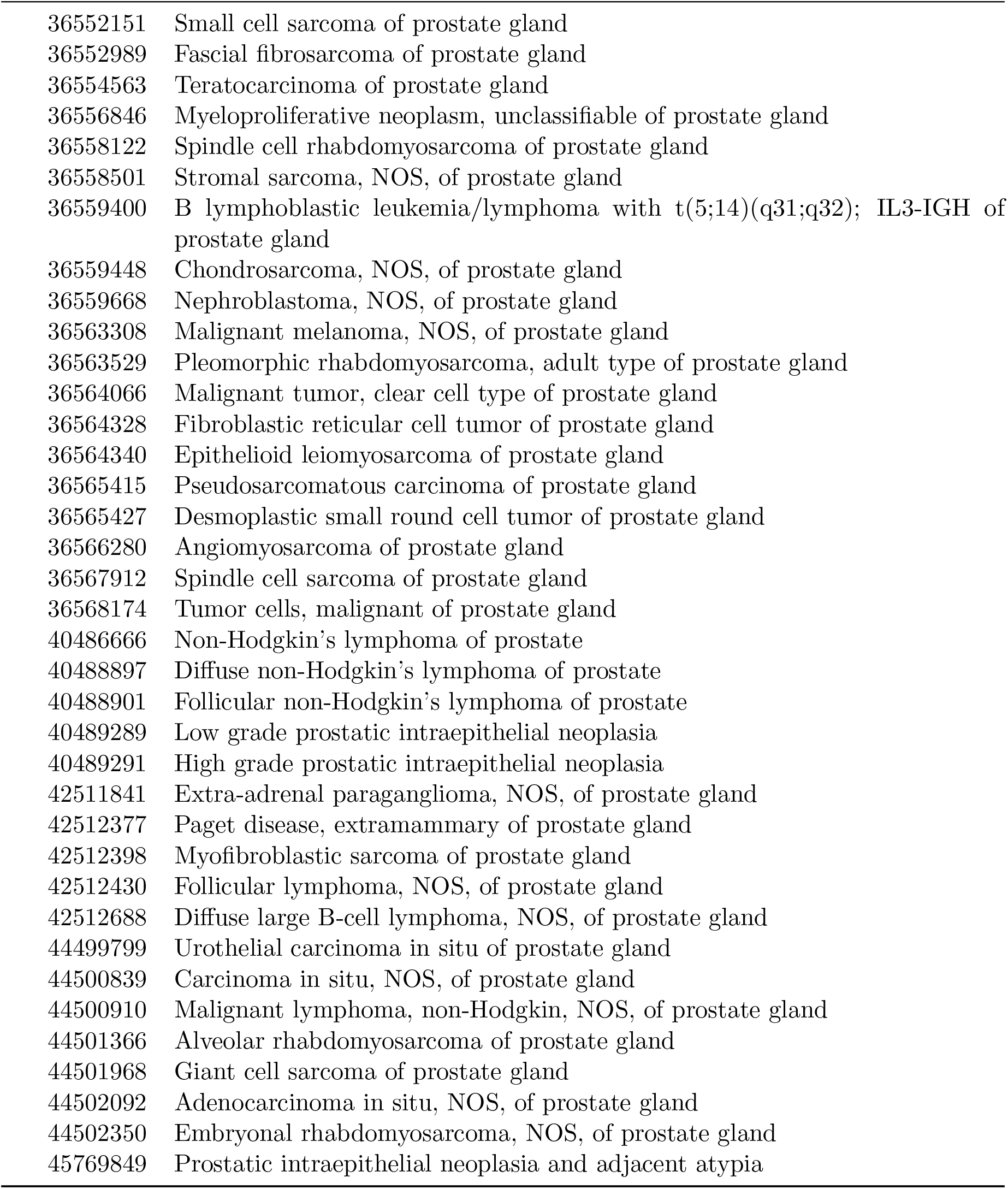
Prostate cancer indeterminate and control-exclusion concepts.

#### D.8 Testicular cancer

**Table D13:** Testicular germ cell tumor and broad-malignancy case concepts.

| Concept ID | Concept name |
| --- | --- |
| 433716 | Primary malignant neoplasm of testis |
| 436054 | Primary malignant neoplasm of undescended testis |
| 619381 | Primary malignant neoplasm of left testis |
| 619382 | Primary malignant neoplasm of right testis |
| 619383 | Bilateral primary malignant neoplasm of testes |
| 4003028 | Primary malignant neoplasm of descended testis |
| 4092524 | Malignant tumor of undescended testis |
| 4095164 | Malignant tumor of ectopic testis |
| 4114339 | Seminoma of testis |
| 4133189 | Mixed seminoma teratoma of testis |
| 4177115 | Malignant tumor of testis |
| 4193703 | Seminoma of descended testis |
| 4197601 | Seminoma of undescended testis |
| 4217083 | Malignant teratoma of undescended testis |
| 4219402 | Malignant teratoma of descended testis |
| 4220146 | Malignant teratoma of testis |
| 4306529 | Seminoma of testis, stage 3 |
| 4309671 | Seminoma of testis, stage 2 |
| 4309811 | Seminoma of testis, stage 1 |
| 4313771 | Seminoma of testis, stage 4 |
| 35610170 | Malignant neoplasm of descended testis |
| 36521552 | Polyembryoma of undescended testis |
| 36524505 | Malignant teratoma, undifferentiated of undescended testis |
| 36525037 | Malignant teratoma, intermediate of testis, NOS |
| 36530105 | Malignant teratoma, trophoblastic of testis, NOS |
| 36532477 | Teratoma with malignant transformation of undescended testis |
| 36533891 | Teratoma with malignant transformation of testis, NOS |
| 36540915 | Malignant teratoma, trophoblastic of undescended testis |
| 36546145 | Polyembryoma of testis, NOS |
| 36547010 | Teratoma with malignant transformation of descended testis |
| 36547066 | Yolk sac tumor, NOS, of undescended testis |
| 36547328 | Polyembryoma of descended testis |
| 36547800 | Malignant teratoma, trophoblastic of descended testis |
| 36549712 | Malignant teratoma, intermediate of undescended testis |
| 36550324 | Malignant teratoma, undifferentiated of testis, NOS |
| 36553786 | Malignant teratoma, intermediate of descended testis |
| 36554633 | Malignant teratoma, undifferentiated of descended testis |
| 36558353 | Seminoma, anaplastic of undescended testis |
| 36568342 | Choriocarcinoma, NOS, of undescended testis |
| 36712742 | Primary seminoma of left testis |
| 36712743 | Primary seminoma of right testis |

Table D13 – continued from previous page
| Concept ID | Concept name |
| --- | --- |
| 36715833 | Primary choriocarcinoma of testis |
| 36717487 | Primary embryonal carcinoma of testis |
| 37016122 | Non-seminomatous germ cell neoplasm of testis |
| 37017381 | Malignant germ cell tumor of testis |
| 44498978 | Choriocarcinoma combined with other germ cell elements of undescended testis |
| 44499767 | Yolk sac tumor, NOS, of descended testis |
| 44500520 | Teratocarcinoma of descended testis |
| 44500690 | Seminoma, anaplastic of testis, NOS |
| 44500691 | Germ cell tumor, nonseminomatous of descended testis |
| 44500898 | Teratocarcinoma of undescended testis |
| 44500899 | Teratocarcinoma of testis, NOS |
| 44500948 | Choriocarcinoma, NOS, of descended testis |
| 44501541 | Seminoma, anaplastic of descended testis |
| 44501817 | Mixed germ cell tumor of descended testis |
| 44502187 | Embryonal carcinoma, NOS, of descended testis |
| 44502561 | Choriocarcinoma combined with other germ cell elements of testis, NOS |
| 44502839 | Choriocarcinoma combined with other germ cell elements of descended testis |
| 44502897 | Embryonal carcinoma, NOS, of undescended testis |
| 44503172 | Yolk sac tumor, NOS, of testis, NOS |
| 44505590 | Germinoma of undescended testis |
| 44505591 | Germ cell tumor, nonseminomatous of undescended testis |
| 44505594 | Mixed germ cell tumor of undescended testis |
| 44505600 | Germinoma of descended testis |
| 44505610 | Germinoma of testis, NOS |

## References

R Alroughani, SF Ahmed, R Behbehani, R Khan, A Thussu, KJ Alexander, A Ashkanani, V Nagarajan, and J Al-Hashel. Increasing prevalence and incidence rates of multiple sclerosis in kuwait. Multiple Sclerosis Journal, 20(5):543–547, 2014.

April W Armstrong, Manan D Mehta, Clayton W Schupp, George C Gondo, Stacie J Bell, and Christopher EM Griffiths. Psoriasis prevalence in adults in the united states. JAMA dermatology, 157(8):940–946, 2021.

Celine Bellenguez, Fahri Kucukali, Iris E Jansen, Luca Kleineidam, Sonia Moreno-Grau, Najaf Amin, Adam C Naj, Rafael Campos-Martin, Benjamin Grenier-Boley, Victor Andrade, et al. New insights into the genetic etiology of alzheimer’s disease and related dementias. Nature genetics, 54 (4):412–436, 2022.

April P Carson, George Howard, Gregory L Burke, Steven Shea, Emily B Levitan, and Paul Muntner. Ethnic differences in hypertension incidence among middle-aged and older adults: the multi-ethnic study of atherosclerosis. Hypertension, 57(6):1101–1107, 2011.

Vincent Chen, Hongyan Ning, Norrina Allen, Kiarri Kershaw, Sadiya Khan, Donald M Lloyd-Jones, and John T Wilkins. Lifetime risks for hypertension by contemporary guidelines in african american and white men and women. JAMA cardiology, 4(5):455–459, 2019.

Xingyu Chen, Fei Wang, Hongqiang Zhao, Jing Hao, Yunga A, Xiong Yang, Tingfeng Xu, Yubo Zhou, Qiuli Chen, Rufan Zhang, et al. Comprehensive benchmarking single and multi ancestry polygenic score methods with the pgs-hub platform. Nature Communications, 17(1):2014, 2026.

Chamlee Cho, Beomsu Kim, Dan Say Kim, Mi Yeong Hwang, Injeong Shim, Minku Song, Yeong Chan Lee, Sang-Hyuk Jung, Sung Kweon Cho, Woong-Yang Park, et al. Large-scale cross-ancestry genome-wide meta-analysis of serum urate. Nature communications, 15(1):3441, 2024.

Siew Eng Choon, Alison K Wright, Christopher EM Griffiths, Kwee Eng Tey, Kit Wan Wong, Yoong Wei Lee, Ushananthiny Suvelayutnan, Jeevitha Mariapun, Darren M Ashcroft, and Global Psoriasis Atlas. Incidence and prevalence of psoriasis in multiethnic johor bahru, malaysia: a population-based cohort study using electronic health data routinely captured in the teleprimary care (tpc®) clinical information system from 2010 to 2020. British Journal of Dermatology, 187 (5):713–721, 2022.

S Cordogan, DB Starr, NR Treff, J Lanchbury, E Goldstein, K Sadeghi, M Burmeister, L Dayani, P Macdonald, JD Keen-Kim, et al. Within-and between-family validation of nine polygenic risk scores developed in 1.5 million individuals: implications for ivf, embryo selection, and reduction in lifetime disease risk. medRxiv, pages 2025–10, 2025.

Nick Dand, Philip E Stuart, John Bowes, David Ellinghaus, Joanne Nititham, Jake R Saklatvala, Maris Teder-Laving, Laurent F Thomas, Tanel Traks, Steffen Uebe, et al. Gwas meta-analysis of psoriasis identifies new susceptibility alleles impacting disease mechanisms and therapeutic targets. Nature Communications, 16(1):2051, 2025.

Anirban Das and Yan Cui. Bridging ancestry gaps in genomic risk prediction with tabular foundation models. Bioinformatics, 42(Supplement 1):btag217, 2026.

Katrina M De Lange, Loukas Moutsianas, James C Lee, Christopher A Lamb, Yang Luo, Nicholas A Kennedy, Luke Jostins, Daniel L Rice, Javier Gutierrez-Achury, Sun-Gou Ji, et al. Genome-wide association study implicates immune activation of multiple integrin genes in inflammatory bowel disease. Nature genetics, 49(2):256–261, 2017.

EADB, EADI, Bonn, ADGC, CHARGE, FinnGen, GERAD, GR@ACE/DEGESCO, and PGC-ALZ. Consensus meta-analysis of genome-wide association studies for alzheimer’s disease and related dementias. Nature Genetics, 58:1214–1225, 2026. doi: 10.1038/s41588-026-02583-1. URL 10.1038/s41588-026-02583-1.

Colin A Ellis, Karen L Oliver, Rebekah V Harris, Ruth Ottman, Ingrid E Scheffer, Heather C Mefford, Michael P Epstein, Samuel F Berkovic, and Melanie Bahlo. Inflation of polygenic risk scores caused by sample overlap and relatedness: Examples of a major risk of bias. The American Journal of Human Genetics, 111(9):1805–1809, 2024.

David S. Friedman, Roger C. W. Wolfs, Benita J. O’Colmain, Barbara E. Klein, Hugh R. Taylor, Shelia West, M. Cristina Leske, Paul Mitchell, Nathan Congdon, John Kempen, and Eye Diseases Prevalence Research Group. Prevalence of open-angle glaucoma among adults in the united states. Archives of Ophthalmology, 122(4):532–538, 2004. doi: 10.1001/archopht.122.4.532. URL https://pmc.ncbi.nlm.nih.gov/articles/PMC2798086/.

Steven Gazal, Hilary K Finucane, Nicholas A Furlotte, Po-Ru Loh, Pier Francesco Palamara, Xuanyao Liu, Armin Schoech, Brendan Bulik-Sullivan, Benjamin M Neale, Alexander Gusev, et al. Linkage disequilibrium–dependent architecture of human complex traits shows action of negative selection. Nature genetics, 49(10):1421–1427, 2017.

Tian Ge, Chia-Yen Chen, Yang Ni, Yen-Chen Anne Feng, and Jordan W Smoller. Polygenic prediction via bayesian regression and continuous shrinkage priors. Nature communications, 10 (1):1776, 2019.

Puya Gharahkhani, Eric Jorgenson, Pirro Hysi, Anthony P Khawaja, Sarah Pendergrass, Xikun Han, Jue Sheng Ong, Alex W Hewitt, Ayellet V Segre, John M Rouhana, et al. Genome-wide meta-analysis identifies 127 open-angle glaucoma loci with consistent effect across ancestries. Nature communications, 12(1):1258, 2021.

Xikun Han, Kaiah Steven, Ayub Qassim, Henry N Marshall, Cameron Bean, Michael Tremeer, Jiyuan An, Owen M Siggs, Puya Gharahkhani, Jamie E Craig, et al. Automated ai labeling of optic nerve head enables insights into cross-ancestry glaucoma risk and genetic discovery in¿ 280,000 images from ukb and clsa. The American Journal of Human Genetics, 108(7):1204–1216, 2021.

Poeya Haydarlou, Daria R Kramarenko, Nobuyuki Enzan, Marie Klevjer, Oliver B Vad, Marre E Corver, Dominic S Zimmerman, Koichi Matsuda, Søren Z Diederichsen, et al. Multi-trait polygenic risk scores improve genomic prediction of atrial fibrillation across diverse ancestries. Nature Communications, 2026.

Michael Hittle, William J Culpepper, Annette Langer-Gould, Ruth Ann Marrie, Gary R Cutter, Wendy E Kaye, Laurie Wagner, Barbara Topol, Nicholas G LaRocca, Lorene M Nelson, et al. Population-based estimates for the prevalence of multiple sclerosis in the united states by race, ethnicity, age, sex, and geographic region. JAMA neurology, 80(7):693–701, 2023.

Laurence J Howe, Michel G Nivard, Tim T Morris, Ailin F Hansen, Humaira Rasheed, Yoonsu Cho, Geetha Chittoor, Rafael Ahlskog, Penelope A Lind, Teemu Palviainen, et al. Within-sibship genome-wide association analyses decrease bias in estimates of direct genetic effects. Nature genetics, 54(5):581–592, 2022.

Seokho Jeong, Manu Shivakumar, Sang-Hyuk Jung, Hong-Hee Won, Kwangsik Nho, Heng Huang, Christos Davatzikos, Andrew J Saykin, Paul M Thompson, Li Shen, et al. Addressing overfitting bias due to sample overlap in polygenic risk scoring. Alzheimer’s & Dementia, 21(4):e70109, 2025.

Jin Jin, Jianan Zhan, Jingning Zhang, Ruzhang Zhao, Jared O’Connell, Yunxuan Jiang, Stella Aslibekyan, Adam Auton, Elizabeth Babalola, Robert K Bell, et al. Mussel: Enhanced bayesian polygenic risk prediction leveraging information across multiple ancestry groups. Cell genomics, 4 (4), 2024.

Linda Kachuri, Nilanjan Chatterjee, Jibril Hirbo, Daniel J Schaid, Iman Martin, Iftikhar J Kullo, Eimear E Kenny, Bogdan Pasaniuc, Polygenic Risk Methods in Diverse Populations (PRIMED) Consortium Methods Working Group Auer Paul L. 20 Conomos Matthew P. 21 Conti David V. 22 23 Ding Yi 24 Wang Ying 19 25 26 Zhang Haoyu 27 28 Zhang Yuji 29, John S Witte, et al. Principles and methods for transferring polygenic risk scores across global populations. Nature Reviews Genetics, 25(1):8–25, 2024.

Gunjeet Kaur, Akashdeep Singh Chauhan, Shankar Prinja, Yot Teerawattananon, Malaisamy Muniyandi, Ashu Rastogi, Gaurav Jyani, Karikalan Nagarajan, PVM Lakshmi, Ankur Gupta, et al. Cost-effectiveness of population-based screening for diabetes and hypertension in india: an economic modelling study. The Lancet Public Health, 7(1):e65–e73, 2022.

Jacob M Keaton, Zoha Kamali, Tian Xie, Ahmad Vaez, Ariel Williams, Slavina B Goleva, Alireza Ani, Evangelos Evangelou, Jacklyn N Hellwege, Loic Yengo, et al. Genome-wide analysis in over 1 million individuals of european ancestry yields improved polygenic risk scores for blood pressure traits. Nature genetics, 56(5):778–791, 2024.

Chang-Fu Kuo, Matthew J Grainge, Lai-Chu See, Kuang-Hui Yu, Shue-Fen Luo, Weiya Zhang, and Michael Doherty. Epidemiology and management of gout in taiwan: a nationwide population study. Arthritis research & therapy, 17(1):13, 2015.

Mitja I Kurki, Juha Karjalainen, Priit Palta, Timo P Sipila, Kati Kristiansson, Kati M Donner, Mary P Reeve, Hannele Laivuori, Mervi Aavikko, Mari A Kaunisto, et al. Finngen provides genetic insights from a well-phenotyped isolated population. Nature, 613(7944):508–518, 2023.

Samuel A Lambert, Laurent Gil, Simon Jupp, Scott C Ritchie, Yu Xu, Annalisa Buniello, Aoife McMahon, Gad Abraham, Michael Chapman, Helen Parkinson, et al. The polygenic score catalog as an open database for reproducibility and systematic evaluation. Nature genetics, 53(4):420–425, 2021.

Sang Hong Lee, Michael E Goddard, Naomi R Wray, and Peter M Visscher. A better coefficient of determination for genetic profile analysis. Genetic epidemiology, 36(3):214–224, 2012.

James D Lewis, Lauren E Parlett, Michele L Jonsson Funk, Colleen Brensinger, Virginia Pate, Qufei Wu, Ghadeer K Dawwas, Alexandra Weiss, Brad D Constant, Maureen McCauley, et al. Incidence, prevalence, and racial and ethnic distribution of inflammatory bowel disease in the united states. Gastroenterology, 165(5):1197–1205, 2023.

Yu Liao, Qian Zhu, Kexin Sun, Rongshou Zheng, Huihong Deng, Lifeng Lin, Ruilin Meng, Wenqiang Wei, and Ye Wang. Trends in the incidence, mortality and lifetime risks of female breast and cervical cancer—guangdong province, china, 2023. China CDC Weekly, 7(28):940, 2025.

Luke R Lloyd-Jones, Jian Zeng, Julia Sidorenko, Loıc Yengo, Gerhard Moser, Kathryn E Kemper, Huanwei Wang, Zhili Zheng, Reedik Magi, Tonu Esko, et al. Improved polygenic prediction by bayesian multiple regression on summary statistics. Nature communications, 10(1):5086, 2019.

Ani Manichaikul, Josyf C Mychaleckyj, Stephen S Rich, Kathy Daly, Michele Sale, and Wei-Min Chen. Robust relationship inference in genome-wide association studies. Bioinformatics, 26(22): 2867–2873, 2010.

Carla Marquez-Luna, Steven Gazal, Po-Ru Loh, Samuel S Kim, Nicholas Furlotte, Adam Auton, and Alkes L Price. Incorporating functional priors improves polygenic prediction accuracy in uk biobank and 23andme data sets. Nature Communications, 12(1):6052, 2021.

Nina Mars, Joni V Lindbohm, Pietro della Briotta Parolo, Elisabeth Widen, Jaakko Kaprio, Aarno Palotie, and Samuli Ripatti. Systematic comparison of family history and polygenic risk across 24 common diseases. The American Journal of Human Genetics, 109(12):2152–2162, 2022.

Barry A Miller, Steven M Scoppa, and Eric J Feuer. Racial/ethnic patterns in lifetime and age-conditional risk estimates for selected cancers. Cancer: Interdisciplinary International Journal of the American Cancer Society, 106(3):670–682, 2006.

Spencer Moore, Ivan Davidson, Jonathan Anomaly, Jeremiah H Li, Mohammad Ahangari, Lauren Moissiy, Michael Christensen, Alexander Strudwick Young, David Stern, and Tobias Wolfram. Development and validation of polygenic scores for within-family prediction of disease risks. medRxiv, pages 2025–08, 2025.

KM Venkat Narayan, James P Boyle, Theodore J Thompson, Stephen W Sorensen, and David F Williamson. Lifetime risk for diabetes mellitus in the united states. Jama, 290(14):1884–1890, 2003.

National Cancer Institute. SEER Cancer Statistics Review, 1975–2018. Technical report, National Cancer Institute, Bethesda, MD, April 2021. URL https://seer.cancer.gov/archive/csr/1975_2018/browse_csr.php?pageSEL=sect_01_table.16&sectionSEL=1. Table 1.16. Based on November 2020 SEER data submission.

Oliver Pain, Kylie P Glanville, Saskia P Hagenaars, Saskia Selzam, Anna E Furtjes, Helena A Gaspar, Jonathan RI Coleman, Kaili Rimfeld, Gerome Breen, Robert Plomin, et al. Evaluation of polygenic prediction methodology within a reference-standardized framework. PLoS genetics, 17(5):e1009021, 2021.

Seunghwan Park and Wonil Chung. Comparative evaluation of statistical learning methods for polygenic prediction in uk biobank. Scientific Reports, 2026.

Florian Prive, Julyan Arbel, and Bjarni J Vilhjalmsson. Ldpred2: better, faster, stronger. Bioinformatics, 36(22-23):5424–5431, 2020.

Florian Prive, Julyan Arbel, Hugues Aschard, and Bjarni J Vilhjalmsson. Identifying and correcting for misspecifications in gwas summary statistics and polygenic scores. Human Genetics and Genomics Advances, 3(4), 2022.

Azra Ramezankhani, Ali Siamak Habibi-Moeini, Seyed Saeed Tamehri Zadeh, Fereidoun Azizi, and Farzad Hadaegh. Effect of family history of diabetes and obesity status on lifetime risk of type 2 diabetes in the iranian population. Journal of Global Health, 12:04068, 2022.

Yunfeng Ruan, Yen-Feng Lin, Yen-Chen Anne Feng, Chia-Yen Chen, Max Lam, Zhenglin Guo, Lin He, Akira Sawa, Alicia R Martin, et al. Improving polygenic prediction in ancestrally diverse populations. Nature genetics, 54(5):573–580, 2022.

Saskia Selzam, Stuart J Ritchie, Jean-Baptiste Pingault, Chandra A Reynolds, Paul F O’Reilly, and Robert Plomin. Comparing within-and between-family polygenic score prediction. The American journal of human genetics, 105(2):351–363, 2019.

Mathias Seviiri, Matthew H Law, Jue-Sheng Ong, Puya Gharahkhani, Pierre Fontanillas, Catherine M Olsen, David C Whiteman, and Stuart MacGregor. A multi-phenotype analysis reveals 19 susceptibility loci for basal cell carcinoma and 15 for squamous cell carcinoma. Nature communications, 13(1):7650, 2022.

Johanna L Smith, Kristjan Norland, Marwan E Hamed, Yue Yu, Jie Na, Ozan Dikilitas, Daniel J Schaid, and Iftikhar J Kullo. Performance of polygenic risk scores for atherosclerotic cardiovascular disease in the all of us program. Circulation: Genomic and Precision Medicine, page e005382, 2026.

Cathie Sudlow, John Gallacher, Naomi Allen, Valerie Beral, Paul Burton, John Danesh, Paul Downey, Paul Elliott, Jane Green, Martin Landray, Bette Liu, Paul Matthews, Giok Ong, Jill Pell, Alan Silman, Alan Young, Tim Sprosen, Tim Peakman, and Rory Collins. UK Biobank: An open access resource for identifying the causes of a wide range of complex diseases of middle and old age. PLoS Medicine, 12(3):e1001779, 2015. doi: 10.1371/journal.pmed.1001779. URL 10.1371/journal.pmed.1001779.

Ken Suzuki, Konstantinos Hatzikotoulas, Lorraine Southam, Henry J Taylor, Xianyong Yin, Kim M Lorenz, Ravi Mandla, Alicia Huerta-Chagoya, Giorgio EM Melloni, Stavroula Kanoni, et al. Genetic drivers of heterogeneity in type 2 diabetes pathophysiology. Nature, 627(8003):347–357, 2024.

The All of Us Research Program Genomics Investigators. Genomic data in the All of Us research program. Nature, 627:340–346, 2024. doi: 10.1038/s41586-023-06957-x. URL 10.1038/s41586-023-06957-x.

Deborah J Thompson, Daniel Wells, Saskia Selzam, Iliana Peneva, Rachel Moore, Kevin Sharp, William A Tarran, Edward J Beard, Fernando Riveros-Mckay, Carla Giner-Delgado, et al. A systematic evaluation of the performance and properties of the uk biobank polygenic risk score (prs) release. Plos one, 19(9):e0307270, 2024.

Kristin Tsuo, Zhuozheng Shi, Tian Ge, Ravi Mandla, Kangcheng Hou, Yi Ding, Bogdan Pasaniuc, Ying Wang, and Alicia R Martin. All of us diversity and scale yield context-dependent improvements in polygenic prediction. Nature Genetics, pages 1–9, 2026.

Emil Uffelmann, Qin Qin Huang, Nchangwi Syntia Munung, Jantina De Vries, Yukinori Okada, Alicia R Martin, Hilary C Martin, Tuuli Lappalainen, and Danielle Posthuma. Genome-wide association studies. Nature reviews methods primers, 1(1):59, 2021.

Anurag Verma, Jennifer E. Huffman, Alex Rodriguez, Mitchell Conery, Molei Liu, et al. Diversity and scale: Genetic architecture of 2068 traits in the VA Million Veteran Program. Science, 385 (6706):eadj1182, 2024. doi: 10.1126/science.adj1182. URL 10.1126/science.adj1182.

Hannah Wand, Samuel A Lambert, Cecelia Tamburro, Michael A Iacocca, Jack W O’Sullivan, Catherine Sillari, Iftikhar J Kullo, Robb Rowley, Jacqueline S Dron, Deanna Brockman, et al. Improving reporting standards for polygenic scores in risk prediction studies. Nature, 591(7849): 211–219, 2021.

Anqi Wang, Jiayi Shen, Alex A Rodriguez, Edward J Saunders, Fei Chen, Rohini Janivara, Burcu F Darst, Xin Sheng, Yili Xu, Alisha J Chou, et al. Characterizing prostate cancer risk through multi-ancestry genome-wide discovery of 187 novel risk variants. Nature genetics, 55(12):2065–2074, 2023.

Zhaoming Wang, Katherine A McGlynn, Ewa Rajpert-De Meyts, D Timothy Bishop, Charles C Chung, Marlene D Dalgaard, Mark H Greene, Ramneek Gupta, Tom Grotmol, Trine B Haugen, et al. Meta-analysis of five genome-wide association studies identifies multiple new loci associated with testicular germ cell tumor. Nature genetics, 49(7):1141–1147, 2017.

Jacob Williams, Tony Chen, Xing Hua, Wendy Wong, Kai Yu, Peter Kraft, Xihao Li, and Haoyu Zhang. Integrating common and rare variants improves polygenic risk prediction across diverse populations. Nature Communications, 2026.

Hao Wu, Paulino Perez-Rodrıguez, Michael Boehnke, Yuehua Cui, Xiaoyu Liang, Ana I Vazquez, and Gustavo de Los Campos. Improving polygenic score prediction for underrepresented groups through transfer learning. Nature Communications, 17(1):1973, 2026.

Leqi Xu, Geyu Zhou, Wei Jiang, Haoyu Zhang, Yikai Dong, Leying Guan, and Hongyu Zhao. Jointprs: A data-adaptive framework for multi-population genetic risk prediction incorporating genetic correlation. Nature Communications, 16(1):3841, 2025.

Leqi Xu, Yikai Dong, Xiaowei Zeng, Zeyu Bian, Geyu Zhou, Leying Guan, and Hongyu Zhao. Mixprs enables multi-population and multi-method polygenic risk scores using summary statistics. Nature Genetics, pages 1–12, 2026.

Chio Yokose, Natalie McCormick, Na Lu, Sruthi Tanikella, Kehuan Lin, Amit D Joshi, Laura M Raffield, Erica Warner, Tony Merriman, John Hsu, et al. Trends in prevalence of gout among us asian adults, 2011-2018. JAMA network open, 6(4):e239501, 2023.

Daigo Yoshida, Tomoyuki Ohara, Jun Hata, Mao Shibata, Yoichiro Hirakawa, Takanori Honda, Yoshihiko Furuta, Emi Oishi, Satoko Sakata, Shigenobu Kanba, et al. Lifetime cumulative incidence of dementia in a community-dwelling elderly population in japan. Neurology, 95(5): e508–e518, 2020.

Alexander I Young, Seyed Moeen Nehzati, Stefania Benonisdottir, Aysu Okbay, Hariharan Jayashankar, Chanwook Lee, David Cesarini, Daniel J Benjamin, Patrick Turley, and Augustine Kong. Mendelian imputation of parental genotypes improves estimates of direct genetic effects. Nature genetics, 54(6):897–905, 2022.

Haoyu Zhang, Thomas U Ahearn, Julie Lecarpentier, Daniel Barnes, Jonathan Beesley, Guanghao Qi, Xia Jiang, Tracy A O’Mara, Ni Zhao, Manjeet K Bolla, et al. Genome-wide association study identifies 32 novel breast cancer susceptibility loci from overall and subtype-specific analyses. Nature genetics, 52(6):572–581, 2020.

Zhili Zheng, Shouye Liu, Julia Sidorenko, Ying Wang, Tian Lin, Loic Yengo, Patrick Turley, Alireza Ani, Rujia Wang, Ilja M Nolte, et al. Leveraging functional genomic annotations and genome coverage to improve polygenic prediction of complex traits within and between ancestries. Nature genetics, 56(5):767–777, 2024.

Geyu Zhou, Tianqi Chen, and Hongyu Zhao. Sdprx: A statistical method for cross-population prediction of complex traits. The American Journal of Human Genetics, 110(1):13–22, 2023.

Wei Zhou, Jonas B Nielsen, Lars G Fritsche, Rounak Dey, Maiken E Gabrielsen, Brooke N Wolford, Jonathon LeFaive, Peter VandeHaar, Sarah A Gagliano, Aliya Gifford, et al. Efficiently controlling for case-control imbalance and sample relatedness in large-scale genetic association studies. Nature genetics, 50(9):1335–1341, 2018.

